# Estimating the new event-free survival

**DOI:** 10.64898/2026.03.25.26349169

**Authors:** Judith Vilsmeier, Maral Saadati, Kaya Miah, Axel Benner, Hartmut Döhner, Jan Beyersmann

## Abstract

**Background:** In acute myeloid leukemia studies, event-free survival (EFS) is defined as time until treatment failure, relapse, or death, whichever occurs first. Since 2020 and 2022, respectively, the US Food and Drug Administration and the European LeukemiaNet recommend analysing treatment failures as day-1 events. This data modification can lead to a potentially large drop in the estimated EFS at day 1. If censoring occurs, the Kaplan-Meier estimator obtained from the recoded data underestimates this drop. Our aim is to obtain an unbiased estimate for EFS as basis for further inference.

**Methods:** We define “event on day 1” as one event type and “‘event after day 1” as a competing event in the original data and use the Aalen-Johansen estimator of the cumulative incidence curve to estimate event-specific transition probabilities, which are combined in one EFS estimate. To analyse effects on day 1 treatment failure and other post-day-1 EFS events separately, a formal link to cure models is established by equating treatment failures with the “cured” proportion in cure model terminology. Additionally, a variance estimator, confidence intervals, confidence bands, and simultaneous testing procedures are derived.

**Results:** Our new estimation method differs from the Kaplan-Meier estimator in settings in which some treatment failures are censored, as in the interim analysis of the AMLSG 09-09 study. If almost no treatment failures are censored, the two estimation methods do not differ. The cure model and simultaneous testing are able to estimate effects on day 1 treatment failure and other post-day-1 EFS events separately and function independently of whether data is modified.

**Conclusions:** The Kaplan-Meier estimator evaluated on the recoded data underestimates the drop at day 1 if treatment failures are censored. With sufficient follow-up, this bias disappears, and results coincide with our novel approach.

## 1 Introduction

In acute myeloid leukemia (AML) studies, commonly considered time-to-event endpoints are overall survival and event-free survival (EFS). EFS is a composite endpoint defined as the time to death, relapse, or treatment failure, whichever occurs first [1, 2]. EFS is becoming increasingly relevant as a primary endpoint as more post-study treatment options become available, potentially affecting the interpretation of overall survival [2]. Additionally, EFS can be assessed earlier than overall survival and with smaller sample sizes.

Treatment failure is assessed if an individual fails to achieve a certain response, e.g., complete remission, by a predetermined landmark [1, 2]. The predetermined landmark often depends on the duration of the induction cycles, which may vary across patients, and affects time to treatment failure [1]. Since 2020 and 2022, respectively, the US Food and Drug Administration (FDA) and the European LeukemiaNet (ELN) recommend recoding treatment failures as events on day 1 of randomisation or the start of treatment rather than at the time of assessment [1, 2]. This results in a data modification in which the event times of treatment failures are shifted from a later time point, the time point of assessment, to an earlier time point, namely day 1. In the following, the term ‘treatment failure’ will always be understood as treatment failure on day 1.

This data modification impacts the estimation of EFS. One concern is that day-1-EFS is determined by observations after day 1, and the question is whether this may bias the analysis [3, 4]. Recoding multiple events as day-1 events results in EFS dropping right at the beginning, with the extent of the drop varying with the amount of tied data. Montesinos et al. [5] reported EFS curves with day-1-drops of around 60% and 80%, respectively. Döhner et al. [6] reported drops of around 10% and 14% in the two treatment groups of the AMLSG 09-09 study. However, only observed treatment failures can be shifted to day 1. Therefore, the Kaplan-Meier estimator (KME) after data modification, evaluated on day 1, only estimates the probability of *observed* treatment failure. If censoring occurs before the landmark, this is not equal to the probability of treatment failure, which is actually of interest [7]. As a consequence, the day-1-drop may be underestimated by the Kaplan-Meier estimator when it is used after data modification to estimate EFS. This potential bias propagates throughout the estimated survival curve. In practice, trials typically aim for sufficient follow-up until the landmark, but some studies may face more pronounced censoring. Additionally, shifting events to day 1 and the resulting day-1-drop leads to EFS now being a mixture of a discrete part (EFS on day 1) and a continuous part (EFS after day 1). This impacts the interpretation of hazard ratios, which are traditionally viewed as comparing hazard functions over time.

Since the Kaplan-Meier estimator may be biased when applied to the modified data, an alternative estimation method is needed. Additionally, methods to analyse effects on the discrete and the continuous parts of EFS separately are required. For this, a competing risks model is defined, that allows to distinguish between events which are recommended to be recoded as day-1 events and other post-day-1 events. In this setting, we derive an estimator for EFS and a variance estimator, used to compute confidence intervals. Additionally, confidence bands can be obtained using the wild bootstrap, which allows for the analysis time to be determined by the number of observed events [8]. This is a common choice in designing trials, but it leads to censoring and event times being dependent. Furthermore, simultaneous testing is used to test whether the day-1-drop and EFS at a later time point differ between two groups. Likewise, mixture cure model methodology is used to analyse the impact of treatment on treatment failures and other EFS-events separately.

To analyse the difference between the new estimation method and the Kaplan-Meier estimator after data modification these methods are applied to the final analysis data of the AMLSG 09-09 study and its interim analysis. The data example will illustrate that the difference between the novel method and the standard analysis on the recoded data will not be more than moderate. However, differences may be more pronounced with both higher censoring and a higher probability of day-1-failure.

## 2 AMLSG 09-09 study

The AMLSG 09-09 study is an open-label, phase 3 study [6]. The analysis population contained 588 patients from 56 hospitals in Germany and Austria (2010-2017). All patients were at least 18 years old, had newly diagnosed *NPM1*-mutated acute myeloid leukaemia according to WHO classification, and an Eastern Cooperative Oncology Group performance status of 0-2. The standard group received idarubicin, cytarabine, etoposide, and all-trans retinoic acid (ATRA) during the induction cycles, and cytarabine and ATRA during the consolidation cycles. The experimental group received gemtuzumab ozogamicin (GO) in addition to the drugs in the standard group. Allocation to the groups was randomised. The interim analysis was planned after the accrual of 276 participants and was conducted with 286 patients. All participants provided written informed consent. The clinical trial was approved by the ethics committees of all sites and is registered with ClinicalTrials.gov(NCT00893399).

## 3 Methods

### 3.1 Competing risks

In a competing risks setting, all individuals start in the initial state 0 and are at risk of transitioning to one of *K* absorbing states. If an individual transitions into state *k* ∈ {1,…, *K*}, they have a type-*k*-event. Let (*X* (*t*))_*t*≥0_ be the competing risks process with *X* (*t*) ∈ {0, 1, …, *K*}, then *T* is the time until a patient reaches an absorbing state, i.e., *T* = inf {*t >* 0 *X* (*t*) ≠ 0}, and the event type is *X* (*T*). The cause-specific hazards are defined as

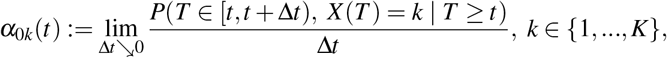

assuming the limits exist. The cumulative cause-specific hazards are defined as

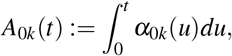

and can be estimated with the Nelson-Aalen estimator

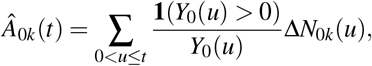

with *Y*_0_(*u*) the number of individuals in state 0 just before event time *u*, Δ*N*_0*k*_(*u*) the number of type-*k*-events observed at *u* and the sum taken over all unique observed event times.

The overall survival *S*(*t*) = P(*T > t*) is the probability that an individual has not had an event of any type until time *t*. It holds that

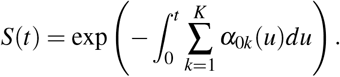

This probability can be estimated with the Kaplan-Meier estimator

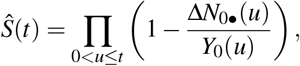

with 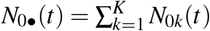 the total number of events in [0, *t*].

The cumulative incidence function (CIF) of event-type *k* is the probability that an individual had a type-*k*-event before or at time *t*, i.e., CIF_*k*_(*t*) = P(*T* ≤ *t, X* (*T*) = *k*). It holds that

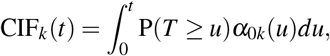

which can be estimated with the Aalen-Johansen estimator

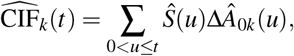

where ΔÂ_0*k*_(*u*) = Δ*N*_0*k*_(*u*)*/Y*_0_(*u*) is the increment of the Nelson-Aalen estimator at *u*. Note that

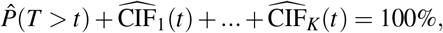

as desired [9].

### 3.2 Event-free survival and ELN/FDA recommendations

Treatment failure is assessed if an individual fails to achieve complete remission by a predetermined landmark [2]. The FDA and ELN recommendations [1, 2] lead to a modified data set in which some event times are shifted from later time points to day 1. Using the Kaplan-Meier estimator to estimate EFS on these modified data may lead to bias, as on day 1 it estimates only the probability of *observed* treatment failure, which is not equal to the probability of treatment failure when censoring occurs before the landmark [7]. Exemplary patient histories and a more detailed explanation of when treatment failure is assessed are available in the appendix.

To derive an alternative method to estimate EFS we define a competing risks model, which can be seen in Figure 1. In this setting, treatment failure is recorded at the time of observation, so there is no data modification.

**Figure 1.**
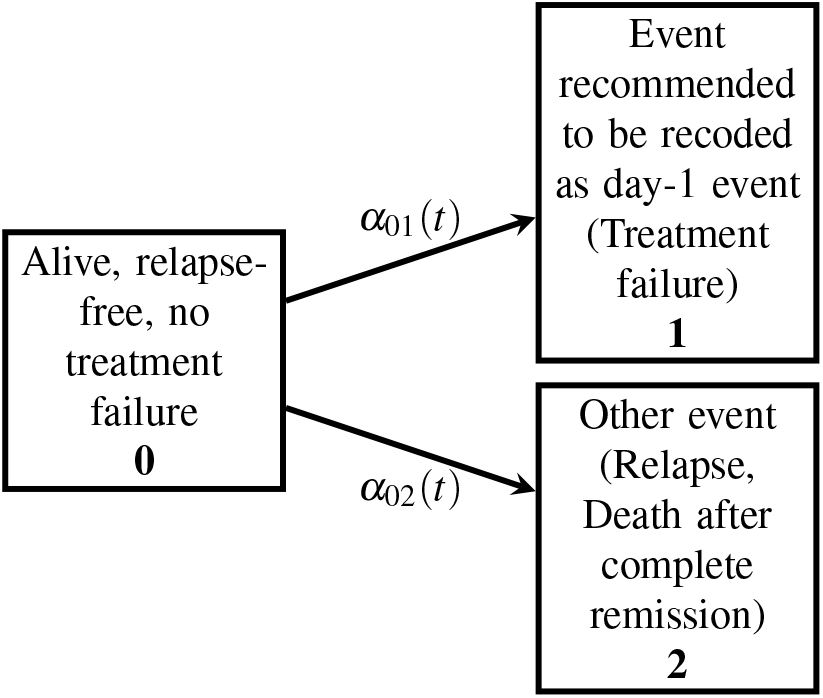
Competing risks model for event-free survival.

Let *T* be the time until assessment of a day-1 event or until occurrence of a post day-1 event, associated with the original data. Let *U* be the EFS time, possibly equal to one (but assessed later). The aim is to estimate EFS(*t*) := *P*(*U > t*). Let *u*_0_ denote the landmark, then it holds that

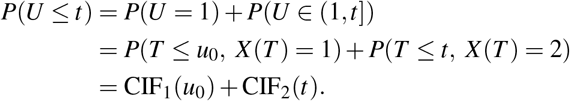

The equality *P*(*U* = 1) = *P*(*T* ≤ *u*_0_, *X* (*T*) = 1) holds, because if an event is recoded as a day-1 event, it is a type-1-event and all type-1-events are observed before or at the landmark. Therefore, it holds that

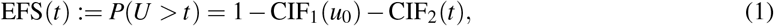

where EFS(1) equals 1 − CIF_1_(*u*_0_), because CIF_2_(1) = 0 by definition. EFS(*t*) can be estimated with the Aalen-Johansen estimator on the original data using the competing risks model.

Using equation (1) one can also derive a variance estimator for 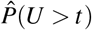:

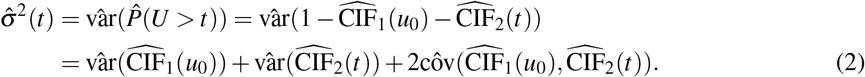

The two different estimated cumulative incidence functions in equation (2) are evaluated at different time points, but one can calculate the covariance using

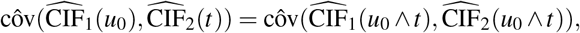

with ∧ denoting the minimum [10]. The variance and covariance estimators are provided in the appendix.

### 3.3 Confidence intervals and confidence bands

The derived variance estimator can be used to obtain point-wise confidence intervals for EFS. The (1 − *α*) ·

100% log-log transformed confidence interval is given by

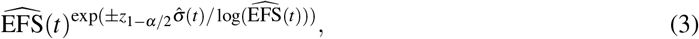

with *z*_1− *α/*2_ the (1 − *α/*2)-quantile of the standard normal distribution and *α* ∈ (0, 1).

Alternatively, one can use resampling methods to obtain confidence intervals. Commonly used is Efron’s bootstrap, i.e., drawing with replacement from the patients, but it has the prerequisite that the data have to be independent and identically distributed (i.i.d.), which is, e.g., not fulfilled in event-driven interim analyses [8]. An alternative is the wild bootstrap [11], which does not have the prerequisite of i.i.d. data and can be easily adapted to linear combinations of Aalen-Johansen estimators as in equation (1).

These resampling methods can also be used to compute simultaneous confidence bands. The difference between confidence intervals and confidence bands is that confidence intervals only cover the true value with probability 1 − *α* in a point-wise manner, i.e., for an arbitrary but fixed time point t, while simultaneous confidence bands cover the entire true EFS curve with probability 1 − *α* over a prespecified interval. Confidence bands can also be obtained for the difference in EFS between two independent groups [11], yielding a Kolmogorov-Smirnov-type test.

### 3.4 Simultaneous testing

Simultaneous testing methods can be used to compare the day-1-drop and EFS at a later time point between two groups. For this, one needs to derive the joint distribution of EFS at time point 1 and a later time point *t >* 1. Let *g* be a group indicator for two independent groups, *g* ∈ {0, 1}. Then, for each group *g*, it holds that

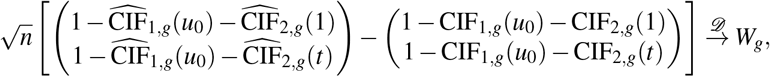

with *W*_*g*_ = (*W*_1,*g*_,*W*_2,*g*_)^*T*^ abivariate Gaussian process with mean (0, 0)^*T*^ . The covariance matrix of *W*_*g*_ is given by

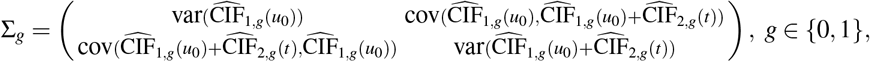

as CIF_2,*g*_(1) = 0 ∀ *g*. Estimators for the elements of Σ_*g*_ can be found in the appendix. To test whether there is a significant difference between groups at time points 1 and *t* simultaneously, one can then use maximum-type tests [12, 13], utilising the independence of the groups.

### 3.5 Link to mixture cure models

To investigate the impact of treatment on treatment failures and other EFS-events separately, one can use cure model methodology by establishing a formal link to mixture cure models. Let *T* and *U* be defined as above. Define Θ as the time until a type-2-event occurs, i.e., relapse/death after complete remission. If an individual has a type-2-event, it holds that *T* = Θ = *U* . If an individual has a type-1-event, *T* is the time point at which the event was observed, while *U* is equal to 1, and Θ is infinity. With these definitions, it holds that

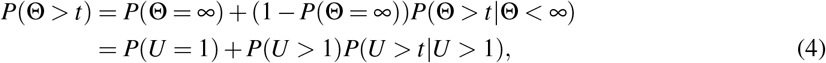

which has the form of a mixture cure model, if one equals the probability of having a type-1-event (treatment failure), with the cured probability in cure model terminology. Of course, day-1-EFS is not a “cure”, but the methodological link is that patients with a day-1-EFS-event do not have an EFS-event afterwards. Therefore, mixture cure models provide a framework to model effects on the treatment failure proportion and relapse-free survival among patients without treatment failure.

For this, let *δ*_*i*_ denote the indicator whether individual *i* had an observed EFS-event, i.e., *δ*_*i*_ = 1 if the individual had a treatment failure, relapsed, or died, and *δ*_*i*_ = 0 if the individual was censored. Additionally, let *s*_*i*_ denote the indicator whether an individual was observed to reach complete remission, i.e., *s*_*i*_ = 1 if complete remission was reached and *s*_*i*_ = 0 if not.

With these definitions, there are four types of individuals: First, individuals who had an observed type-2-event, i.e, those who died or relapsed after reaching complete remission. For them, it holds that *δ*_*i*_ · *s*_*i*_ = 1. Second, individuals who had an observed type-1-event, i.e., treatment failure. They did not reach complete remission, so for them *δ*_*i*_ · (1 − *s*_*i*_) = 1 holds. The remaining individuals were censored. Some individuals are censored after reaching complete remission and can only have an (unobserved) type-2-event. For them, it holds that (1 − *δ*_*i*_) · *s*_*i*_ = 1. The others are censored before reaching complete remission and can have an unobserved event of either type. For them, (1 − *δ*_*i*_) · (1 − *s*_*i*_) = 1 holds.

Let 𝒪 = {(*t*_*i*_, *δ*_*i*_, *s*_*i*_, **x**_*i*_, **z**_*i*_), *i* = 1, 2, …, *n*}be the observed data of n individuals, with *t*_*i*_ the individual exit time and **x**_*i*_ and **z**_*i*_ covariate vectors. Additionally, let *π*(*x*) := *P*(*U* = 1 | **X** = **x**) denote the probability of treatment failure on day 1 given covariates **x** and *S*(*t* | **z**) := *P*(*U > t* | *U >* 1, **Z** = **z**) the probability of not having a type-2-event until time *t* conditional on no treatment failure and covariates **z**. Then the observed likelihood is

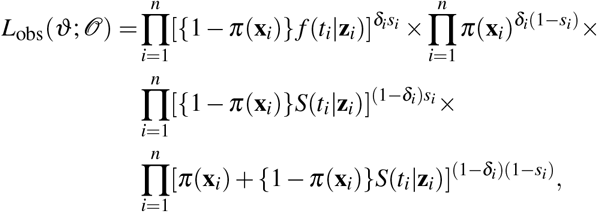

with *ϑ* a vector containing all model parameters and *f* (*t* **z**) the density of *S*(*t* **z**). The notation of **x** and **z** allows for different covariates in the models. Nevertheless, we use the treatment group as the only covariate in both models in addition to an intercept term, i.e., **x** = **z**. For the estimation of the treatment failure probability, a logistic regression model is chosen, i.e.,

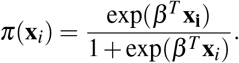

To model the relapse-free survival of individuals without treatment failure, a Weibull model is used, i.e.,

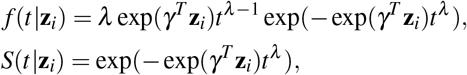

with *λ >* 0 a shape parameter. With these choices it holds that ϑ = (*β, γ, λ*) [14, 15]. The model is fitted using a parametric likelihood approach [14].

## 4 Results

### 4.1 Comparison of estimation methods

First, we compare the day-1-drop in EFS estimated with the Kaplan-Meier estimator after data modification and the cumulative incidence function of type-1-events estimated with the Aalen-Johansen estimator. At interim, 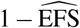 (1) is 0.147 (experimental) and 0.098 (standard) using the new method compared to 0.137 and 0.088, respectively, using the Kaplan-Meier estimator after data modification. Therefore, there is a difference of about 0.01 between 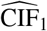(*u*_0_) and 1 KME(1) in both groups. In the final analysis 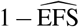(1) is 0.098 in the standard group with both estimation methods, as well as 0.141 in the experimental group using the new method and 0.140 using the Kaplan-Meier estimator after data modification. So in the final analysis, there is no practically difference between estimation methods. A figure comparing 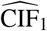(*u*_0_) and 1-KME(1) is available in the appendix.

The reason that there is a difference in the interim analysis but not in the final analysis is that in the final analysis there is almost no censoring before the landmark (2 observations in the standard group, 6 in the experimental group), while in the interim analysis more observations are being censored (23 observations in the standard group, 19 in the experimental group). This shows that the Kaplan-Meier estimator after data modification only estimates the probability of observed treatment failure, which is not equal to the probability of treatment failure if observations are censored before the landmark, which leads to an underestimation of the day-1-drop.

Figure 2 illustrates that this difference, caused by the underestimation of the day-1-event-probability by the Kaplan-Meier estimator, propagates through the EFS curves at interim, but there is no visible difference in the final analysis. Figure 3 shows EFS estimated with the Aalen-Johansen estimator together with log-log transformed 95% confidence intervals obtained using equation (3) and simultaneous log-log transformed 95% confidence bands obtained with the wild bootstrap. Since the confidence bands include the entire true curve, they are wider than the confidence intervals, which cover the true values in a point-wise manner. The differences between groups with confidence bands are shown in Figure 4 for the interim and final analysis. In both analyses, the zero line is included in the confidence bands, so there are no significant differences between groups.

**Figure 2.**
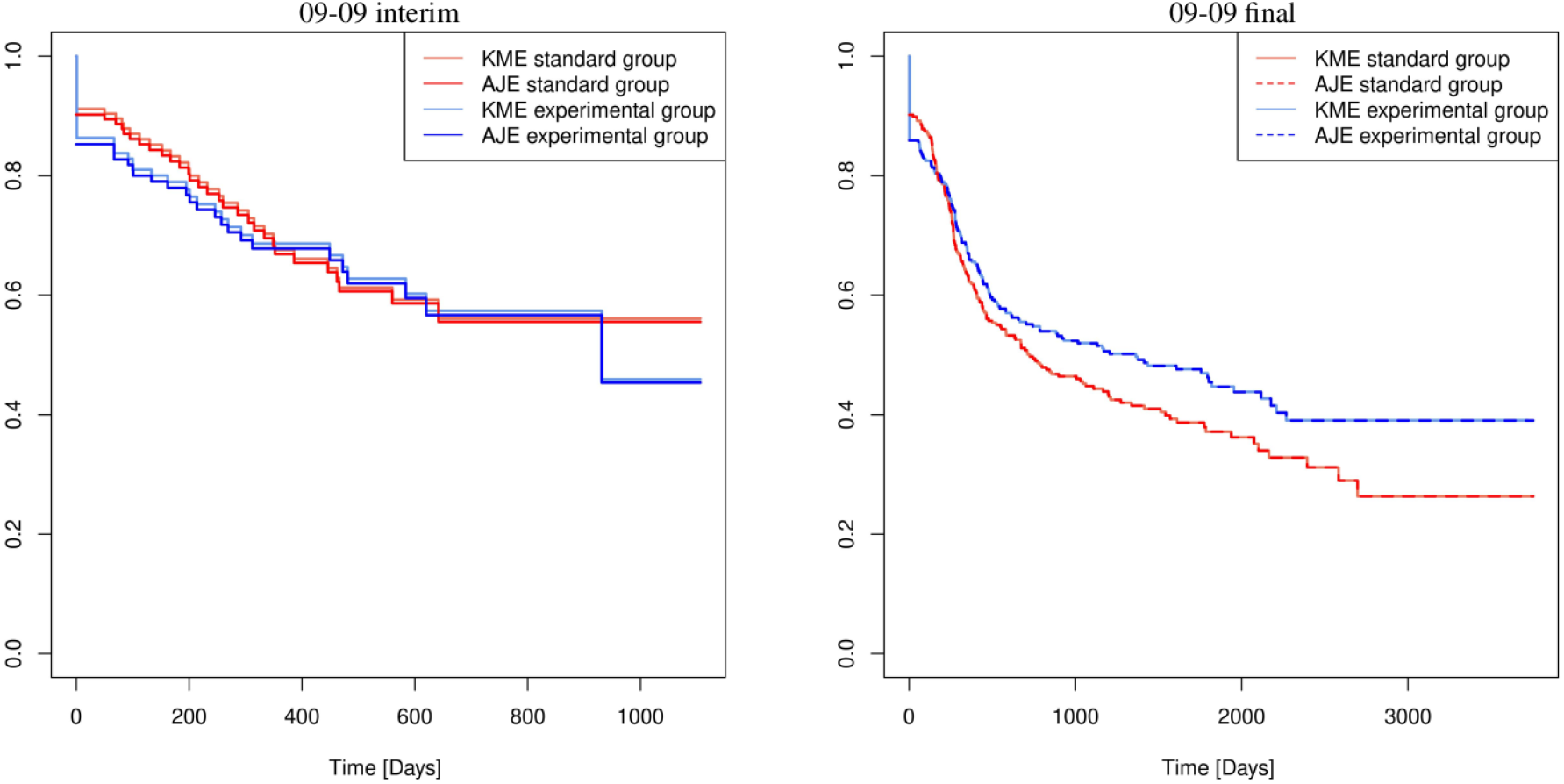
Comparison between event-free survival estimated with the Aalen-Johansen estimator in the competing risk setting and event-free survival estimated with the Kaplan-Meier estimator on the modified data in the final analysis of the 09-09 study (right panel) and its interim analysis (left panel).

**Figure 3.**
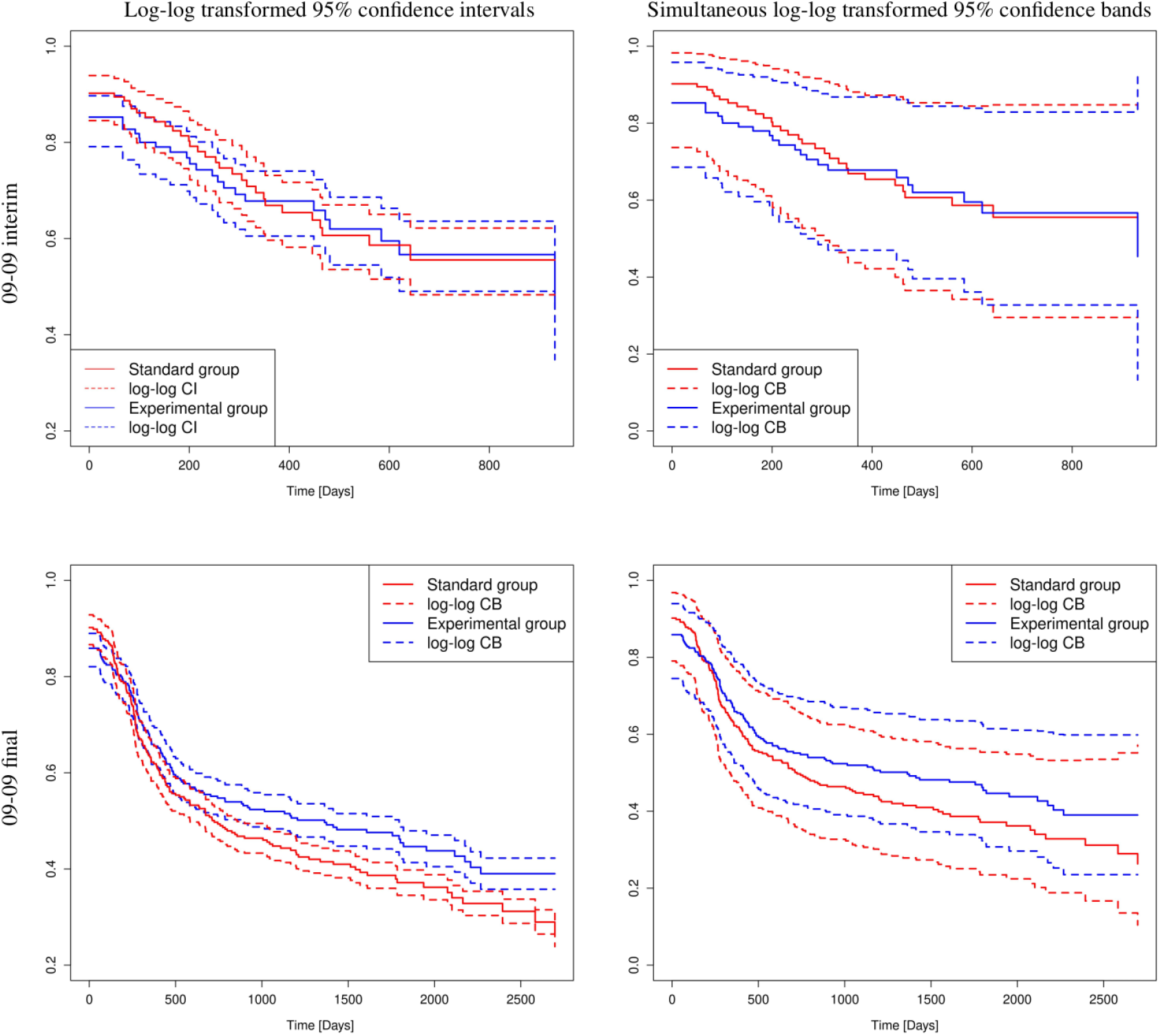
Log-log transformed 95% confidence intervals for event-free survival estimated with the Aalen-Johansen estimator computed using equation (3) (left column) and simultaneous log-log transformed 95% confidence bands obtained with the wild bootstrap (right column) for the final analysis of the 09-09 study (bottom row) and its interim analysis (top row).

**Figure 4.**
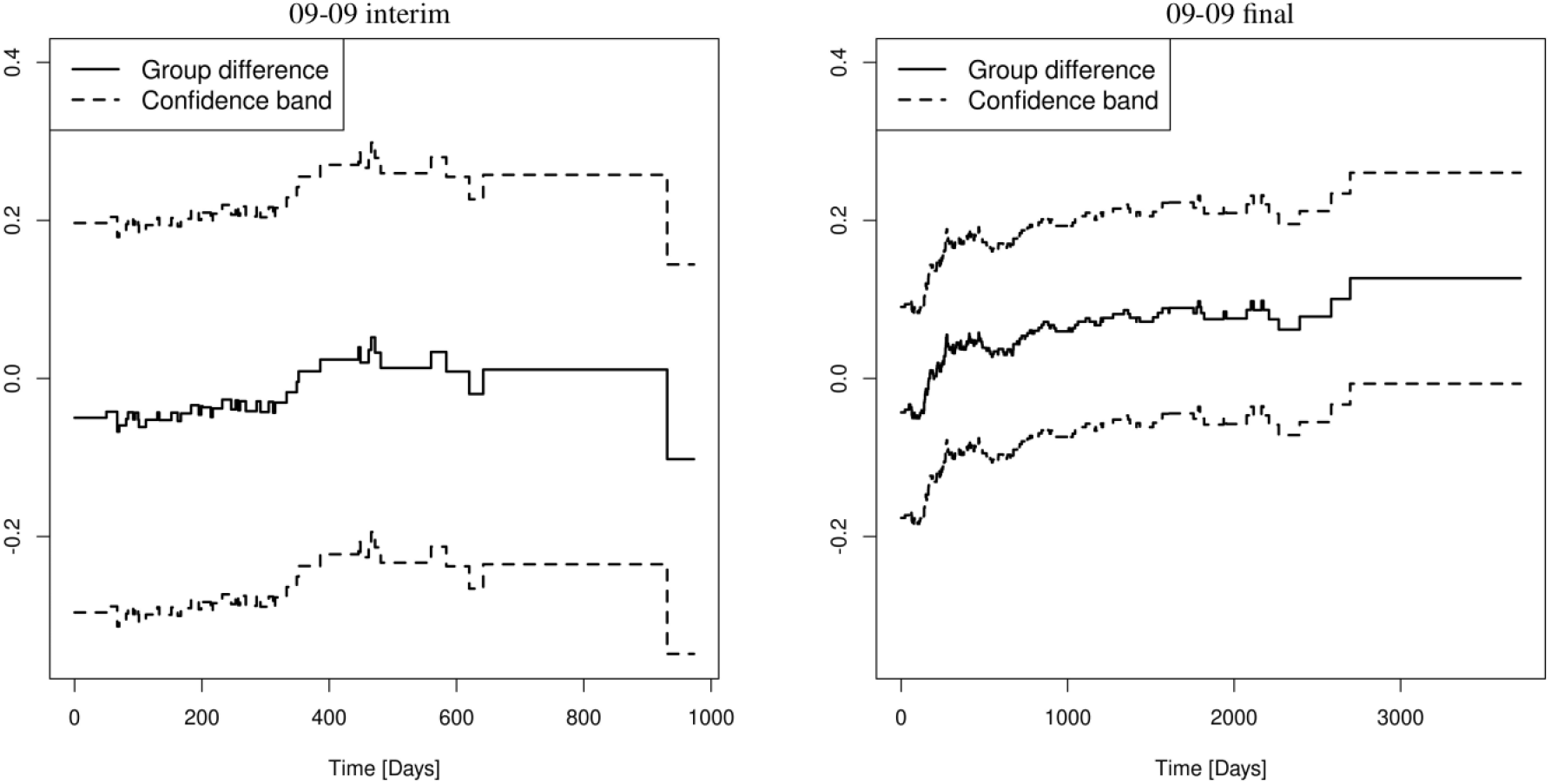
Difference between groups (experimental - standard) and its 95% confidence band in the final analysis of the 09-09 study (right panel) and its interim analysis (left panel).

### 4.2 Simultaneous testing and mixture cure model

The results of the simultaneous testing of EFS at day 1 and a later time point are shown in Table 1. In the final analysis of the 09-09 study, *t* = 10 years (3652 days) was chosen as the later time point, and *t* = 2 years (730 days) was chosen in the interim analysis. At interim, neither the difference at day 1 nor the difference at 2 years is significant. In the final analysis, differences are still non-significant, but there is a signal for 10-year-EFS with a p-value of 8.9%.

**Table 1.** Results of the simultaneous testing of the day-1-drop and event-free survival at a later time point *t*.

| Study | t (days) | Group difference (experimental - standard) | adjusted P-value |
| --- | --- | --- | --- |
| 09-09 interim | 1 | -0.04969 | 0.3715 |
|  | 730 | 0.01127 | 0.9894 |
| 09-09 final | 1 | -0.04292 | 0.1962 |
|  | 3652 | 0.12693 | 0.0883 |

The results of the mixture cure model are shown in Table 2. Note that here, the analysis of the hazard ratios is conditional on an intercurrent event (no treatment failure). In the final analysis, there is no significant evidence that the treatment failure proportion differs between groups. For the relapse-free survival among patients without treatment failure, there is a significantly lower instantaneous risk of dying or relapsing in the experimental group. In the interim analysis, there is no significant evidence that the treatment failure proportion or the relapse-free survival without treatment failure differ between groups.

**Table 2.**
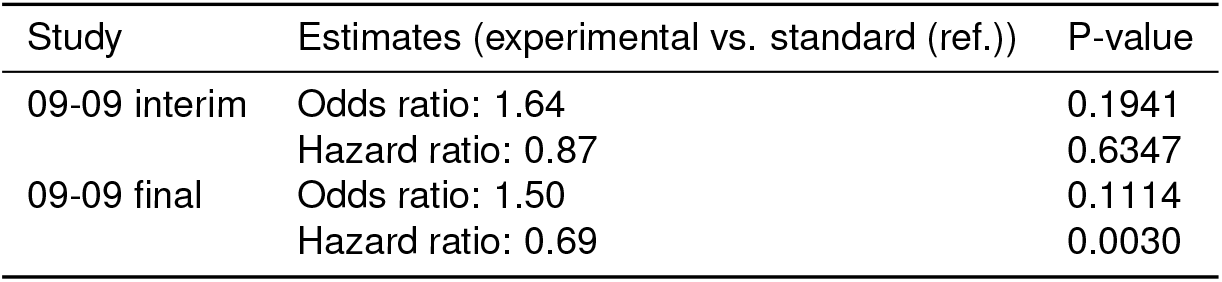
Results of a mixture cure model, in which proportion of treatment failures is modeled with a logistic regression model and the relapse-free survival among patients without treatment failure is modeled with a Weibull model.

The fact that the odds ratios are quite different from the hazard ratios indicates that a Cox or log-rank analysis of the complete EFS curve would be questionable. The results suggest that treatment may have a beneficial effect on EFS despite a higher, non-significant day-1-probability in the final analysis of the 09-09 study. A section on predicting EFS using the cure model is available in the appendix, along with a sensitivity analysis of the results in Table 2 regarding model fit.

## 5 Discussion

The FDA and ELN recommend recoding treatment failures as events on day 1 after randomisation or start of treatment [1, 2]. However, this data modification may lead to bias in the presence of censoring. To avoid this, we derived an estimand - rather than a data modification - and an unbiased estimation method for EFS. To distinguish between events recommended to be recoded as day-1 events and other events, a competing risks setting was outlined in which EFS can be estimated with the Aalen-Johansen estimator. This estimation method provides an unbiased estimate for EFS in which treatment failures are accounted for in a way that is consistent with the idea of the recommendations.

Our data examples from the AMLSG 09-09 study demonstrated that the Kaplan-Meier estimator after data modification underestimates the day-1-drop if censoring occurs before the landmark. This underestimation does not occur when EFS is estimated with the Aalen-Johansen estimator in the competing risks setting. If no censoring occurs, both methods give the same results. Of course, the differences between the two estimation methods in our example data are quite moderate, but the difference could be more pronounced in the case of a larger day-1-drop, as in [5], or more pronounced censoring before the landmark.

Another consequence of the recommendations and the resulting day-1-drop is that EFS is now a mixture of a discrete part and a continuous part, and that the concept of hazard ratios is questionable. Confidence intervals can be computed using the estimated variance of our EFS estimator for pointwise comparisons. Alternatively, resampling methods can be used to obtain simultaneous confidence bands. For this purpose, the wild bootstrap can be used, which can be easily applied to EFS estimated with the Aalen-Johansen estimator and, unlike Efron’s bootstrap, also works with event-driven censoring.

Additionally, we used maximum-type tests to simultaneously test the difference between groups at day 1 and a later time point. If one is not only interested in group comparisons of EFS at day 1 and a later time point, but in the impact of treatment on treatment failures and other EFS-events separately, one can use cure model methodology. We established a formal link to mixture cure models by equating the treatment failure proportion with the cured proportion of a cure model. These methods are able to estimate effects on day 1 treatment failure and other post-day-1 EFS events separately and function independently of whether data is modified. The fact that the odds ratios estimated by the cure model in the example data are quite different from the hazard ratios indicates that a Cox or log-rank analysis of the complete EFS curve would be questionable.

Overall, the FDA and ELN recommendations cannot simply be dismissed, as there are medical reasons underlying the question of the timing of treatment failures. The recommendations have motivated the methodological development in this paper, accounting for the fact that assessment of day-1-EFS may be unobserved due to censoring. The methods used in this paper are based on well-established methodology and implemented in standard statistical software, e.g., R, and can be easily utilised. Only the cure model methodology has to be adapted to our observed likelihood. An example code is provided in the supplementary material. In studies with low censoring before the landmark, modifying the data and using the Kaplan-Meier estimator to estimate EFS works and gives unbiased results.

## Declarations

## Acknowledgments

The authors thank the AMLSG Clinical Trials Office and Daniela Späth for providing the data used in this paper.

## Data availability

This clinical trial data can be requested (by contacting aml.sekretariat@uniklinik ulm.de) by qualified researchers who perform rigorous, independent research; data will be provided after the review and approval of a research proposal and statistical analysis plan and execution of a data sharing agreement. Data will be accessible for 12 months, with possible extensions considered.

## Appendix

### A.1 Exemplary patient histories

To implement the recommendations of the FDA and ELN, one must decide which events constitute treatment failures and which do not. The recommendations state that treatment failure is assessed if an individual fails to achieve a certain response, e.g., complete remission by a predetermined landmark [1, 2]. Also assessed with treatment failure are individuals who die before they can be evaluated for response [2]. To decide which events are considered treatment failures, one has to check the following: If a patient has an EFS-event before the landmark, one has to check whether the patient achieved complete remission before the event occurred or not. If complete remission was achieved, the event is not a treatment failure and therefore a type-2-event. If complete remission was not achieved, the event is labeled as a treatment failure and recommended to be recoded as a day-1 event (type-1-event). If a patient is observed until the landmark and does not have an EFS-event until then, the future course of action again depends on whether the patient has achieved complete remission. If the patient did not achieve complete remission before the landmark, they are assessed with refractory disease/treatment failure at this time point, and the event time is recommended to be recoded as day 1. If complete remission was achieved, the patient continues to be followed up, and any event that may occur later is always of type 2.

Figure A1 shows exemplary patient histories. In the Figure, patients who were censored are omitted as censored observations are never observed treatment failures. Patient D dies before achieving complete remission, and the event is therefore of type 1, i.e., a treatment failure. Patient C has no event before the landmark but fails to achieve complete remission. Because of this, this patient is then assessed with refractory disease/treatment failure at the landmark. Patients A and B die or relapse after achieving complete remission. Their event is therefore of type 2 regardless of whether the event occurs before or after the landmark. Here it becomes clear that treatment failures can only be observed up to the landmark, not afterwards, which justifies *P*(*U* = 1) = *P*(*T* ≤ *u*_0_, *X* (*T*) = 1) in the “Event-free survival and ELN/FDA recommendations” section in the main manuscript.

**Figure A1.**
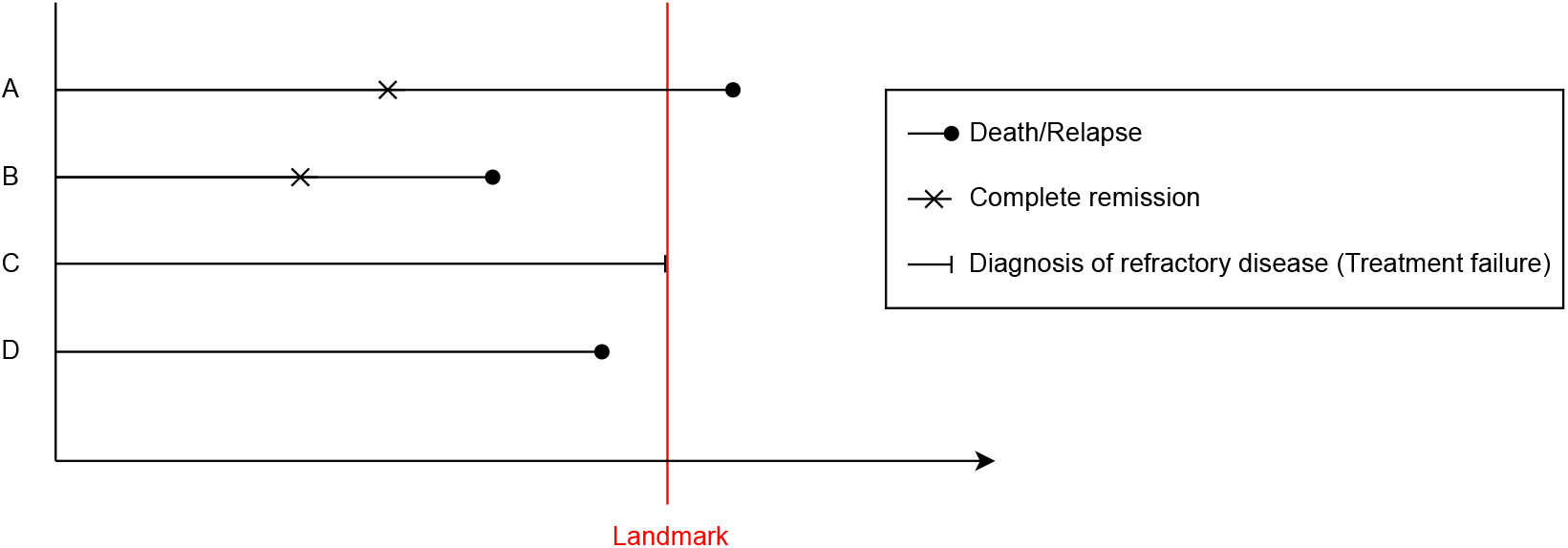
Exemplary patient histories.

### A.2 Variance and covariance estimators

For the variance and covariance estimators, we need to extend the notation from the “Competing risks” section in the main manuscript. Define the transition probability P_0*k*_(*s, t*) as the probability that an individual who is in the initial state 0 at time *s* will be in state *k* at time *t*, i.e., P_0*k*_(*s, t*) = P(*X* (*t*) = *k* | *X* (*s*) = 0), *k* ∈ {0, …, *K*}, *s* ≤ *t*. Then it holds that P(*T > t*) = P_00_(0, *t*), i.e., that the overall survival is equal to the probability that an individual who is in state 0 at time 0 will still be in state 0 at a later time *t*. Additionally, for the cumulative incidence functions it holds that CIF_*k*_(*t*) = P_0*k*_(0, *t*), *k* = 1, …, *K*. Let *Y*_0_(*t*) denote the number of individuals in state 0 just before time *t* and *N*_0*k*_(*t*) the counting process which counts the number of type-*k*-events in [0, *t*]. With these definitions, and *J*_0_(*t*) := **1**(*Y*_0_(*t*) *>* 0), it holds for the variance that

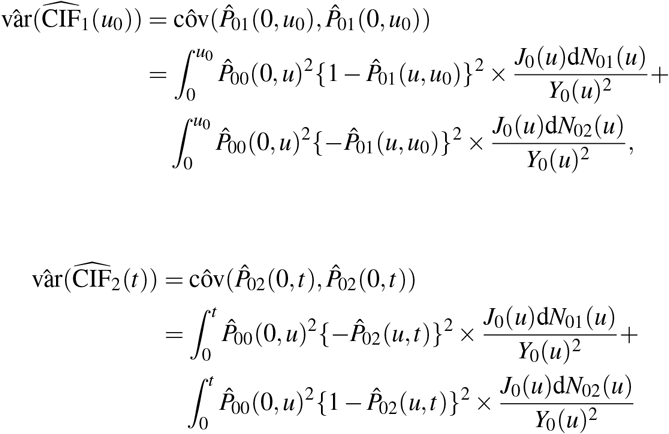

and for the covariance

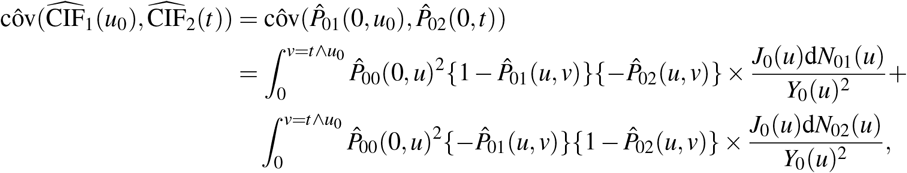

with ∧ denoting the minimum. With these estimators, one can compute 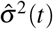 as in equation (2) in the main manuscript. Additionally, one can compute 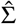, an estimator for the covariance matrix Σ in the “Simultaneous testing” section in the main manuscript, using 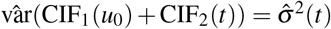 and

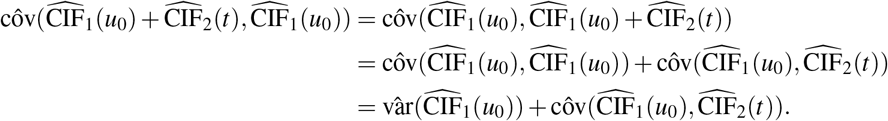

### A.3 Comparison of estimated day-1-EFS

Figure A2 shows the estimated CIF and EFS for the final analysis of the 09-09 study and its interim analysis. Observations which were censored before the landmark are marked by ticks in the plots of the estimated CIFs. The aim is to compare the plateaus of the CIFs (Figure A2, left column) with the day-1-increase of the KMEs (right column).

**Figure A2.**
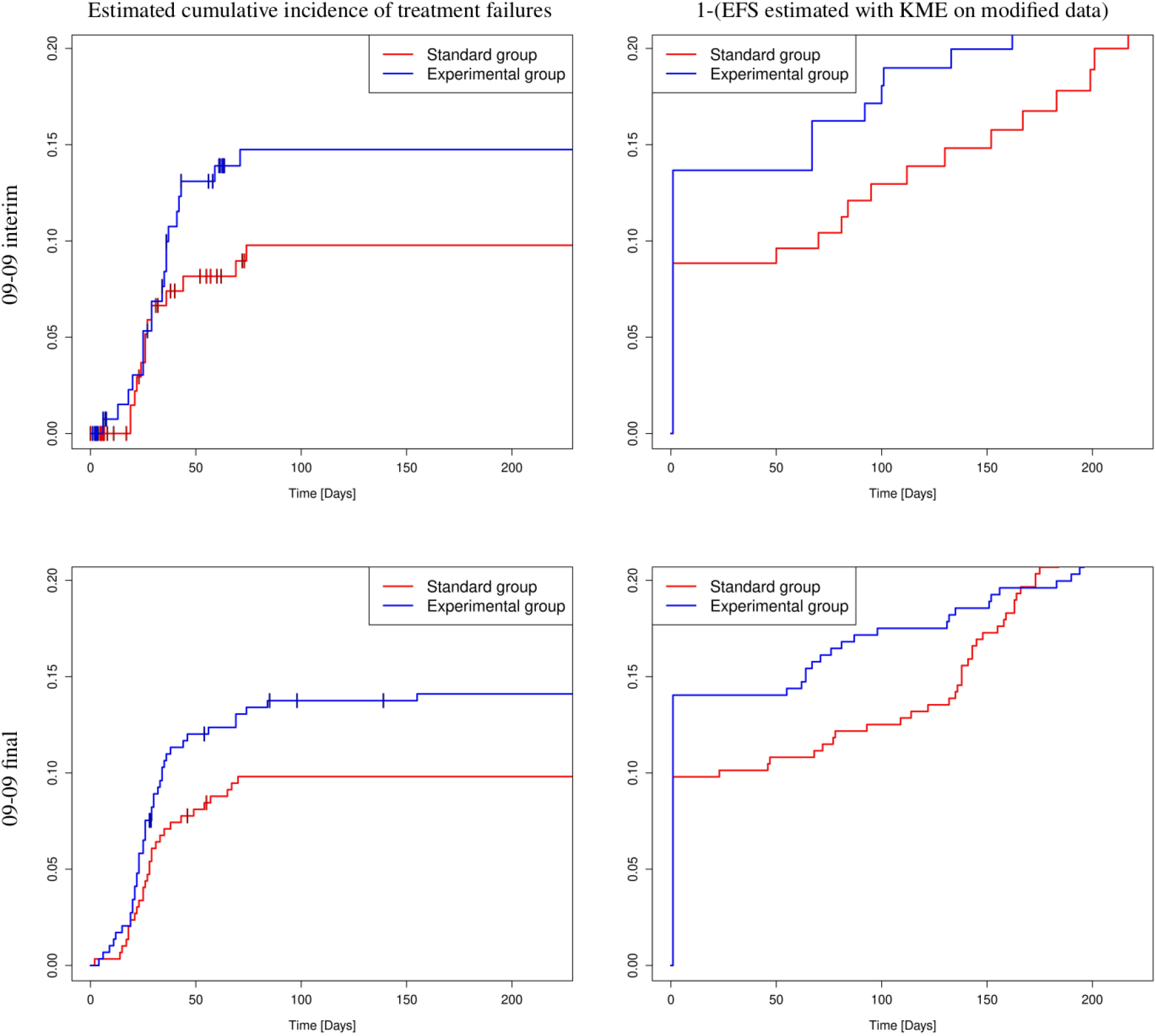
The estimated cumulative incidence function of treatment failures (left column) and 1 minus the event-free survival estimated with the Kaplan-Meier estimator on the modified data (right column) for the final analysis of the 09-09 study (bottom row) and its interim analysis (top row). Observations which were censored before the landmark are marked by ticks in the plot of the estimated cumulative incidence of treatment failures.

The estimated cumulative incidence functions all reach a plateau, after which they do not change anymore. This is because no type-1-events can be observed after the landmark. At interim, one can see a difference between the plateaus of the CIFs and the day-1-increase of the KMEs. In the final analysis there is no visible difference between estimation methods in both groups. One can also see, that in the interim analysis more observations are being censored before the landmark than in the final analysis.

### A.4 Prediction with the cure model

Having a look at equation (4) in the main manuscript, one can see that it holds that

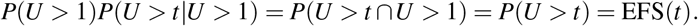

Therefore, one can use the parameters estimated by the cure model to derive a predicted event-free survival by plugging the estimated parameters 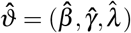 and covariates **x** and **z** into {1 − *π*(**x**)}*S*(*t*_*i*_|**z**).

The results of the prediction can be seen in Figure A3. At interim, predicted EFS is mostly within the 95% confidence interval of the estimated EFS, only at the end, where only a few patients remain under observation, the prediction is lower. In the final analysis, on the other hand, the predicted EFS does not really fit the estimated EFS and is outside the 95% confidence interval. However, the deviation of the predicted EFS from the estimated EFS tends in the same direction in both groups, so the estimates of the odds ratio and hazard ratio in the “Simultaneous testing and mixture cure model” section in the main manuscript should not be too much affected by the not-so-good fit, as we illustrate next.

To improve prediction, the Weibull model can be extended to a piecewise Weibull model with two cutpoints:

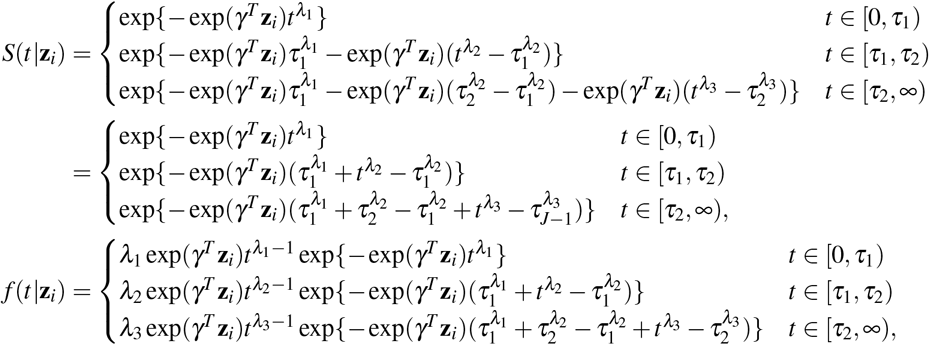

**Figure A3.**
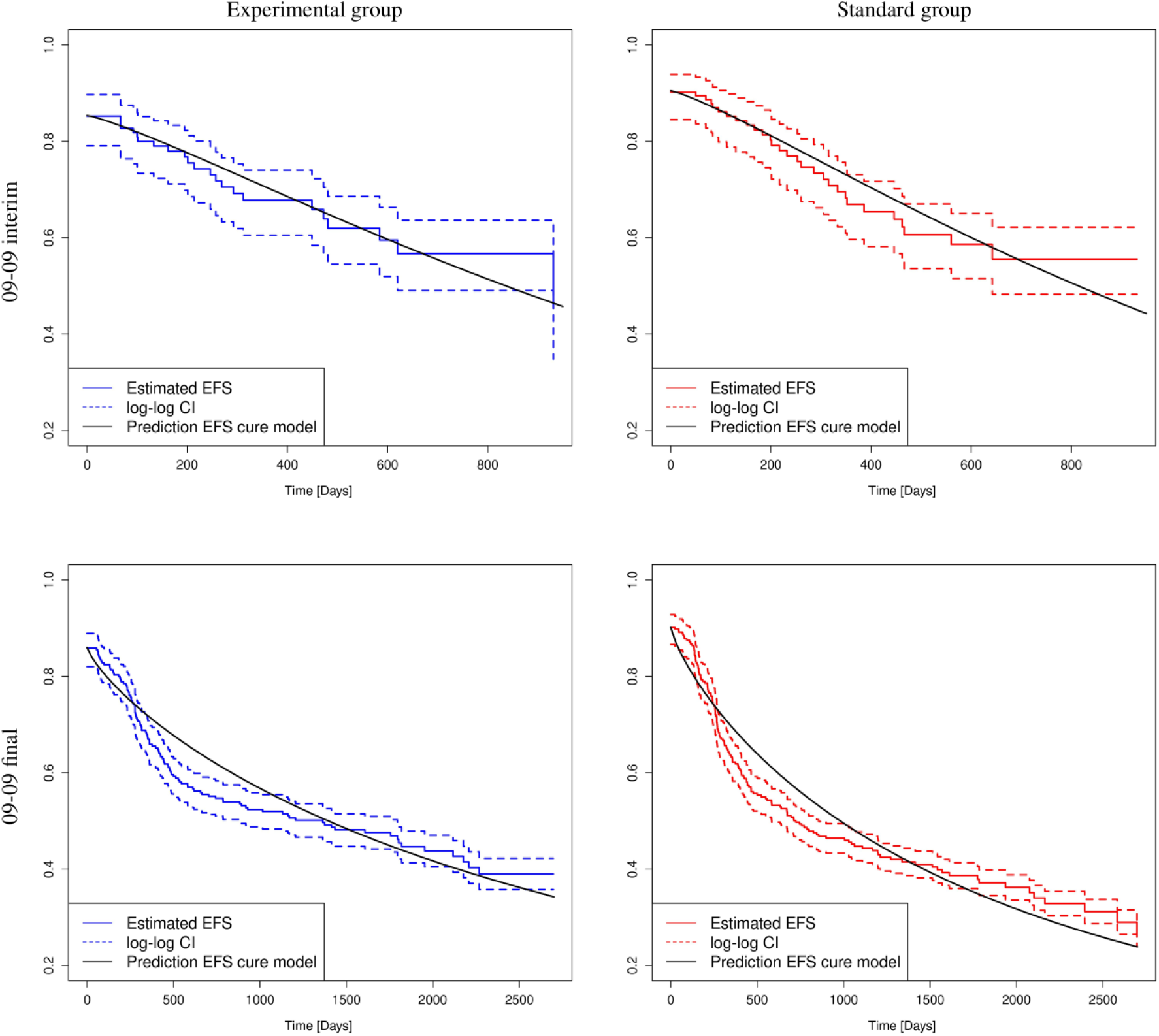
Predicted event-free survival calculated with the parameters estimated by a logistic Weibull cure model and event-free survival estimated with the Aalen-Johansen estimator with log-log transformed 95% confidence intervals in the experimental group (left column) and standard group (right column) for the final analysis of the 09-09 study (bottom row) and its interim analysis (top row).

So for this cure model it holds that ϑ = (*β, γ, λ*_1_, *λ*_2_, *λ*_3_). Note that only the shape varies between time intervals, while the parameters *β* and *γ* stay the same. So, one still assumes proportional hazards, i.e., a constant hazard ratio, with this model.

Results of the prediction using the logistic piecewise Weibull cure model are shown in Figure A4. Here, the cutpoints *τ*_1_ = 100 days and *τ*_2_ = 350 days were chosen for the interim analysis and *τ*_1_ = 200 days and *τ*_2_ = 500 days were chosen for the final analysis. One can see that the EFS predicted with the piecewise cure model fits the estimated EFS better in both analyses and also lies within the 95% confidence intervals.

**Figure A4.**
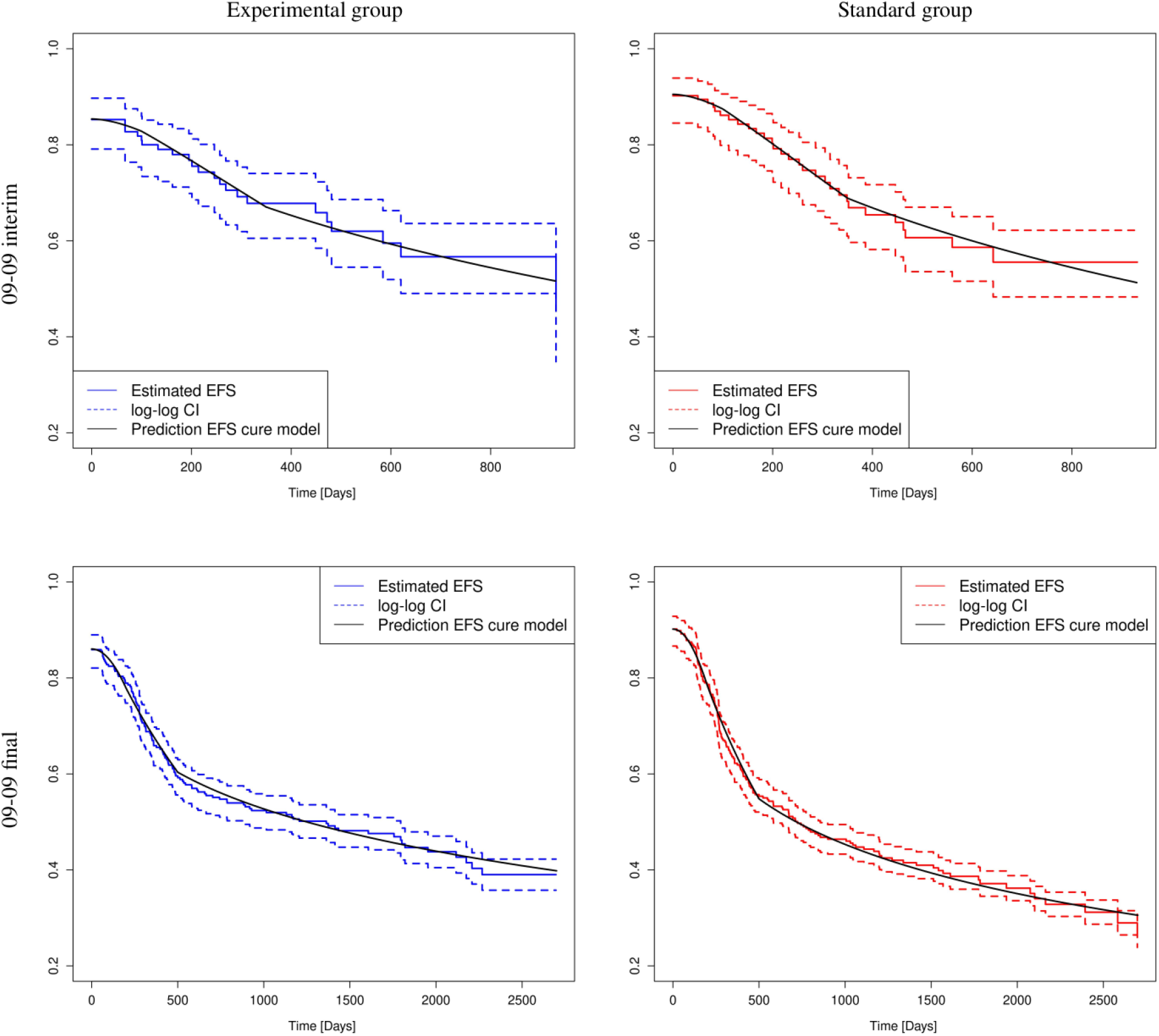
Predicted event-free survival calculated with the parameters estimated by a logistic piecewise Weibull cure model with two cutpoints and event-free survival estimated with the Aalen-Johansen estimator with log-log transformed 95% confidence intervals in the experimental group (left column) and standard group (right column) for the final analysis of the 09-09 study (bottom row) and its interim analysis (top row).

With this improved model, the odds ratio and hazard ratio were estimated again. The results can be seen in Table A1. The estimated odds ratios and hazard ratios are very similar to those estimated in the “Simultaneous testing and mixture cure model” section in the main manuscript. The p-values also do not change much either. So, the simpler logistic Weibull cure model still provides valid estimates of odds ratios and hazard ratios even thought it does not fit the estimated EFS as well as the logistic piecewise Weibull cure model.

**Table A1.** Results of a mixture cure model, in which proportion of treatment failures is modeled with a logistic regression model and the relapse-free survival among patients without treatment failure is modeled with a piecewise Weibull model with two cutpoints.

| Study | Estimates (experimental vs. standard (ref.)) | P-value |
| --- | --- | --- |
| 09-09 interim | Odds ratio: 1.65 | 0.2030 |
|  | Hazard ratio: 0.87 | 0.7231 |
| 09-09 final | Odds ratio: 1.49 | 0.1130 |
|  | Hazard ratio: 0.70 | 0.0063 |

