## Supplementary material for "Estimating the new event-free survival": Code supplement: EFS.R

```
rm(list=ls())
library(survival)
library(etm)
library(multcomp)

setwd(dir = "/home/judith/Nextcloud/Latex/EFS/Code supplement/")

load('./data_example.RData')
# id: patient id
# cr = s in paper: = 1 if complete remission was reached, = 0 if not
# EFS_event = delta in paper: = 1 if any EFS-event (relapse/death/treatment failure), 0 if censored
# event_type: event type in competing risks model, event_type = 1 is a treatment failure (recommended to be recorded on day 1), event_type = 2 is an EFS-event not recommended to be recorded on day 1 (relapse/death after CR)
# time: event time points or time points at which the patients were last observed in days 
# time_yr: event time points or time points at which the patients were last observed in years 
# time_modified: time after data modification (Treatment failure recorded at day 1)
# group: standard or experimental group

###############################################################################################################################################
########## Estimation EFS ##########

##### EFS with KME on modified data #####
efs_kme <- survfit(Surv(time_modified,EFS_event)~group, data=data_example)

#################################################
##### EFS with AJE in competing risks model #####
source("./Functions/est_EFS.R")
efs_aje <- est_EFS(data=data_example, time=time, competing_event=event_type, landmark = 180, group=group, conf.lev=0.95)

############################################################################
##### Comparison of estimation methods (KME modified vs AJE comp risks) #####
## Comparison 1-KME(1) with CIF_1(landmark) 
# Standard group
1-summary(efs_kme)$surv[summary(efs_kme)$strata == "group=standard"][1]
# 1-KME(1) = 0.14
efs_aje[["group standard"]]$CIF_1_landmark
# CIF_1(landmark) = 0.151

# Experimental group
1-summary(efs_kme)$surv[summary(efs_kme)$strata == "group=experimental"][1]
# 1-KME(1) = 0.163
efs_aje[["group experimental"]]$CIF_1_landmark
# CIF_1(landmark) = 0.175

## Comparison KME(1) with EFS_AJE(1) 
# Standard group
summary(efs_kme)$surv[summary(efs_kme)$strata == "group=standard"][1]
# KME(1) = 0.86
efs_aje[["group standard"]]$efs[1]
# EFS_AJE(1) = 0.849

# Experimental group
summary(efs_kme)$surv[summary(efs_kme)$strata == "group=experimental"][1]
# KME(1) = 0.837
efs_aje[["group experimental"]]$efs[1]
# EFS_AJE(1) = 0.825

## Comparison KME(t) with 1-CIF_1(landmark)-CIF_2(t)
# Plot 
dev.new()
plot(efs_kme,col=c("cornflowerblue","coral2"), lwd = 2,xlab="Time [Days] ", ylab="", main = "EFS")
lines(efs_aje[["group standard"]]$time, efs_aje[["group standard"]]$efs, type = "s", col = "red", lwd = 2, lty = 2)
lines(efs_aje[["group experimental"]]$times, efs_aje[["group experimental"]]$efs, type = "s", col = "blue", lwd = 2, lty = 2)
legend("topright",lty=c(1,2,1,2),col=c("coral2","red","cornflowerblue","blue"),legend=c("KME standard group","AJE standard group","KME experimental group","AJE experimental group"))

###################################################################
##### EFS (AJE) with log-log transformed confidence intervals ####
# Plot EFS in standard group with CI
dev.new()
plot(efs_aje[["group standard"]]$time, efs_aje[["group standard"]]$efs, type = "s", col="red", lwd = 2, ylim=c(0,1), xlab="Time [Days] ", ylab="", main = "Standard group: EFS with log-log CI")
lines(efs_aje[["group standard"]]$time, efs_aje[["group standard"]]$ci_loglog[1,], type = "s", col = "black", lwd = 2, lty = 2)
lines(efs_aje[["group standard"]]$time, efs_aje[["group standard"]]$ci_loglog[2,], type = "s", col = "black", lwd = 2, lty = 2)
legend("topright",lty=c(1,2),col=c("red","black"),legend=c("EFS","log-log CI"))

# Plot EFS in experimental group with CI
dev.new()
plot(efs_aje[["group experimental"]]$times, efs_aje[["group experimental"]]$efs, type = "s", col="blue", lwd = 2, ylim=c(0,1), xlab="Time [Days] ", ylab="", main = "Experimental group: EFS with log-log CI")
lines(efs_aje[["group experimental"]]$times, efs_aje[["group experimental"]]$ci_loglog[1,], type = "s", col = "black", lwd = 2, lty = 2)
lines(efs_aje[["group experimental"]]$times, efs_aje[["group experimental"]]$ci_loglog[2,], type = "s", col = "black", lwd = 2, lty = 2)
legend("topright",lty=c(1,2),col=c("blue","black"),legend=c("EFS","log-log CI"))

# Plot both groups together 
dev.new()
plot(efs_aje[["group standard"]]$time, efs_aje[["group standard"]]$efs, type = "s", lwd = 2, col="red", xlab="Time [Days] ", ylab="", ylim=c(0.2,1), main = "EFS with log-log CI")
lines(efs_aje[["group standard"]]$time, efs_aje[["group standard"]]$ci_loglog[1,], type = "s", col = "red", lwd = 2, lty = 2)
lines(efs_aje[["group standard"]]$time, efs_aje[["group standard"]]$ci_loglog[2,], type = "s", col = "red", lwd = 2, lty = 2)
lines(efs_aje[["group experimental"]]$times, efs_aje[["group experimental"]]$efs, type = "s", col="blue", lwd = 2)#
lines(efs_aje[["group experimental"]]$times, efs_aje[["group experimental"]]$ci_loglog[1,], type = "s", col = "blue", lwd = 2, lty = 2)
lines(efs_aje[["group experimental"]]$times, efs_aje[["group experimental"]]$ci_loglog[2,], type = "s", col = "blue", lwd = 2, lty = 2)
legend("topright",lty=c(1,2,1,2),col=c("red","red","blue","blue"),legend=c("Standard group","log-log CI","Experimental group","log-log CI"), lwd = c(2,2,2,2))


###############################################################################################################################################
########## Wild bootstrap and confidence bands ##########
noit <- 1000
t.start <- 0
alpha <- 0.05

#Data
# A special data format is needed for WB 
data.etm_stand <- efs_aje[["group standard"]]$data
data.etm_exp <- efs_aje[["group experimental"]]$data

##########################
##### Wild bootstrap #####
source("./Functions/Wild_bootstrap_and_CB/WB_efs.R")

## Wild Bootstrap for each group
# Standard group
landmark_stand <- efs_aje[["group standard"]]$landmark
set.seed(34567)
# Takes some time
WB_AJE_stand <- WB_efs(data.etm_stand, landmark_stand, noit)

# Experimental group
landmark_exp <- efs_aje[["group experimental"]]$landmark
set.seed(23456)
# Takes some time
WB_AJE_exp <- WB_efs(data.etm_exp, landmark_exp, noit)

###############################################################
##### EFS (AJE) with log-log transformed confidence bands #####
source("./Functions/Wild_bootstrap_and_CB/CB_efs.R")

## Compute confidence bands for each group
# Standard group
n_stand <- length(unique(data.etm_stand$id))
# t.end = last event timepoint in group
t.end_stand <- max(data.etm_stand$exit[data.etm_stand$to != "cens"])
efs_stand <- efs_aje[["group standard"]]$efs
times_stand <- efs_aje[["group standard"]]$times
CB_efs_stand <- CB_efs(efs_stand, times_stand, WB_AJE_stand, n_stand, alpha, t.start, t.end_stand)

# Experimental group
n_exp <- length(unique(data.etm_exp$id))
# t.end = last event timepoint in group
t.end_exp <- max(data.etm_exp$exit[data.etm_exp$to != "cens"])
efs_exp<- efs_aje[["group experimental"]]$efs
times_exp <- efs_aje[["group experimental"]]$times
CB_efs_exp <- CB_efs(efs_exp, times_exp, WB_AJE_exp, n_exp, alpha, t.start, t.end_exp)

## Plot (CB starts at first event time, so time point zero has to be removed from times vector when plotting the confidence band)
dev.new()
plot(efs_aje[["group standard"]]$times, efs_aje[["group standard"]]$efs, type = "s", ylim = c(0, 1), xlab = "Time [Days]", ylab = "", col = "red", lwd = 2, main = "EFS with log-log CB")
lines(efs_aje[["group standard"]]$times[-1], CB_efs_stand[,1], type = "s", col = "red", lwd = 2, lty=2)
lines(efs_aje[["group standard"]]$times[-1], CB_efs_stand[,2], type = "s", col = "red", lwd = 2,lty=2)
lines(efs_aje[["group experimental"]]$times, efs_aje[["group experimental"]]$efs, type = "s", col = "blue", lwd = 2)
lines(efs_aje[["group experimental"]]$times[-1], CB_efs_exp[,1], type = "s", col = "blue", lwd = 2, lty=2)
lines(efs_aje[["group experimental"]]$times[-1], CB_efs_exp[,2], type = "s", col = "blue", lwd = 2,lty=2)
legend("bottomleft", lty=c(1,2,1,2),col=c("red","red","blue","blue"), lwd = 2, legend = c("Standard group","log-log CB","Experimental group","log-log CB"), cex=1.2)


#############################################################
##### Linear confidence bands for EFS-curve-differences #####
source("./Functions/Wild_bootstrap_and_CB/CB_linear_diff.R")

## Compute confidence band for group difference (experimental-standard)
t.end <- max(t.end_stand,t.end_exp)
CB_diff <- CB_linear_diff(data.etm_exp, data.etm_stand, WB_AJE_exp, WB_AJE_stand, alpha, t.start, t.end) 

## Plot
dev.new()
plot(CB_diff$times, CB_diff$est, type = "s", xlab = "Time [Days]", ylab = "", ylim = c(-0.5, 0.7), lwd = 2)
lines(CB_diff$times, CB_diff$CB.lower, type = "s", lwd = 2, lty=2)
lines(CB_diff$times, CB_diff$CB.upper, type = "s", lwd = 2,lty=2)
legend("topleft", lty=c(1,2), lwd = 2, legend = c("Group difference","log-log CB"), cex=1.2)


###############################################################################################################################################
########## Cure model ##########
##### Mixture cure model (logistic + Weibull) #####
source("./Functions/Cure_model/log_weib_cure.R")

# does not work if event times get to big, therefore variable time_year is used here instead of time
cure_mod <- log_weib_cure(data=data_example, time=time_year, group=group, delta=EFS_event, s=cr) 
cure_mod$logistic 
cure_mod$weibull 


##################################################################################################
##### Piecewise mixture cure model (logistic + piecewise Weibull with two cutpoints) #####
source("./Functions/Cure_model/piecew_log_weib_cure_twoCuts.R")

# (Cutpoints at 100 days and 500 days)
# Takes some time
set.seed(12345)
# does not work if event times get to big, therefore variable time_year is used here instead of time
piecew_cure_mod <- piecew_log_weib_cure_twoCuts(data=data_example, time=time_year, group=group, delta=EFS_event, s=cr, cutpoints = c(0.27378508,1.36892539))
piecew_cure_mod$logistic
piecew_cure_mod$weibull

###############################################################################################################################################
########## Simultaneous testing ##########
source("./Functions/Simultaneous_testing/simultaneous_testing.R")

# (t = 2 years = 730 days)
simul_test <- simul_test_efs(data=data_example, time=time, competing_event=event_type, group=group, landmark = 155, t = 730, conf.lev=0.95)
simul_test$tab
```
