## Supplementary material for "Estimating the new event-free survival": Code supplement: piecew_log_weib_cure_twoCuts.R

```
piecew_log_weib_cure_twoCuts <- function(data, time, group, delta, s, cutpoints){
  ## ------------------------------------------------------------------------------------------
  ## Author: Judith Vilsmeier
  ## ------------------------------------------------------------------------------------------
  ## Description: Computes a piecewise logistic Weibull cure model with a group variable as the  
  ##              only covariate
  ## ------------------------------------------------------------------------------------------
  ## Required Packages: 
  ## ------------------------------------------------------------------------------------------
  ## Input:      data: dataset with one row per patient
  ##             time: name of the column in data which contains the time points at which the 
  ##                   patients were last observed
  ##            group: name of the column in data which contains the group
  ##            delta: indicator whether an individual had an observed EFS-event (of any type)
  ##                s: indicator whether an individual reached complete remission
  ##        cutpoints: vector containing the two cutpoints for the piecewise cure model
  ## ------------------------------------------------------------------------------------------
  ## Output: A list containing the estimated parameters of the cure model, tables containing 
  ##         the group parameter, OR/HR, and p-value of the logistic model and the Weibull 
  ##         model and tables containing likelihood ratio tests of the group parameters
  ## ------------------------------------------------------------------------------------------
  
  if(missing(data)) stop("Argument 'data' is missing with no default")
  if(missing(time)) stop("Argument 'time' is missing with no default")
  if(missing(group)) stop("Argument 'group' is missing with no default")
  if(missing(delta)) stop("Argument 'delta' is missing with no default")
  if(missing(s)) stop("Argument 's' is missing with no default")
  if(missing(cutpoints)) stop("Argument 'cutpoints' is missing with no default")
  if(!is.data.frame(data)) stop("Argument 'data' must be a data.frame")
  
  call <- match.call()
  time <- with(data,get(call$time))
  group_factor <- as.factor(with(data,get(call$group)))
  #Transform group to a numerical vector containing 0,1,...
  group <- as.numeric(group_factor)-1 
  delta <- with(data,get(call$delta))
  s <- with(data,get(call$s))
  
  if(!is.numeric(time)) stop("Argument 'time' must be a numerical vector.")
  if(!all(group%in%c(0,1))) stop("Argument 'group' must only contain two groups.")
  if(all(group==0) | all(group==1)) stop("Only one group defined in argument 'group'.")
  if(!is.numeric(delta)) stop("Argument 'delta' must be a numerical vector.") 
  if(!all(delta%in%c(0,1))) stop("Argument 'delta' is must be a vector with entries 0 or 1.")
  if(!is.numeric(s)) stop("Argument 's' must be a numerical vector.") 
  if(!all(s%in%c(0,1))) stop("Argument 's' must be a vector with entries 0 or 1.") 
  if(!is.numeric(cutpoints)) stop("Argument 'cutpoints' must be a numerical vector.") 
  if(length(cutpoints) != 2) stop("Argument 'cutpoints' must be of lenght two.")
  
  # Fit full Model
  init_model_full <- c(0,0,0,0,0,0,0)
  names(init_model_full) <- c("beta0", "beta1", "gamma0", "gamma1", "log_shape1", "log_shape2", "log_shape3")
  opt_model_full <- optim(par = init_model_full, fn = lklh.logit.piecew.weib.twoCuts, method = "SANN", control = list(maxit = 1000000), group=group, time=time, delta=delta, s=s, cutpoints=cutpoints)
  
  logistic <- data.frame(X = "group=1", coef = c(opt_model_full$par[2]), exp_coef = c(exp(opt_model_full$par[2])))
  weibull <- data.frame(X = "group=1", coef = c(opt_model_full$par[4]), exp_coef = c(exp(opt_model_full$par[4])))
 
  # Fit model without beta1
  init_model_wo_beta1 <- c(0,0,0,0,0,0)
  names(init_model_wo_beta1) <- c("beta0", "gamma0", "gamma1", "log_shape1", "log_shape2", "log_shape3")
  opt_model_wo_beta1 <- optim(par = init_model_wo_beta1, fn = lklh.model2.ratioTest.beta1.piecew.weib.twoCuts, method = "SANN", control = list(maxit = 1000000), group=group, time=time, delta=delta, s=s, cutpoints=cutpoints)
  
  # Likelihood ratio test for beta 1
  lrt_beta1 <- matrix(rep(NA, 10), ncol = 5)
  colnames(lrt_beta1) <- c("#Df", "LogLik", "Df", "Chisq", "Pr(>Chisq)")
  rownames(lrt_beta1) <- c("Full model", "Model w/o beta1") 
  
  lrt_beta1[, 1] <- c(length(unlist(opt_model_full$par)), length(unlist(opt_model_wo_beta1$par)))
  lrt_beta1[, 2] <- c(-opt_model_full$value, -opt_model_wo_beta1$value)
  lrt_beta1[, 3] <- c(NA, abs(lrt_beta1[2, 1] - lrt_beta1[1, 1]))
  lrt_beta1[, 4] <- c(NA, 2 * abs(lrt_beta1[2, 2] - lrt_beta1[1, 2]))
  lrt_beta1[, 5] <- c(NA, stats::pchisq(lrt_beta1[2, 4], df = lrt_beta1[2, 3], lower.tail=FALSE)) 
  
  logistic$p_value <- lrt_beta1[2,5]
  
  # Fit model without gamma1
  init_model_wo_gamma1 <- c(0,0,0,0,0,0)
  names(init_model_wo_gamma1) <- c("beta0", "beta1", "gamma0", "log_shape1", "log_shape2", "log_shape3")
  opt_model_wo_gamma1 <- optim(par = init_model_wo_gamma1, fn = lklh.model2.ratioTest.gamma1.piecew.weib.twoCuts, method = "SANN", control = list(maxit = 1000000), group=group, time=time, delta=delta, s=s, cutpoints=cutpoints)
  
  # Likelihood ratio test for gamma1
  lrt_gamma1 <- matrix(rep(NA, 10), ncol = 5)
  colnames(lrt_gamma1) <- c("#Df", "LogLik", "Df", "Chisq", "Pr(>Chisq)")
  rownames(lrt_gamma1) <- c("Full model", "Model w/o gamma1") 
  
  lrt_gamma1[, 1] <- c(length(unlist(opt_model_full$par)), length(unlist(opt_model_wo_gamma1$par)))
  lrt_gamma1[, 2] <- c(-opt_model_full$value, -opt_model_wo_gamma1$value)
  lrt_gamma1[, 3] <- c(NA, abs(lrt_gamma1[2, 1] - lrt_gamma1[1, 1]))
  lrt_gamma1[, 4] <- c(NA, 2 * abs(lrt_gamma1[2, 2] - lrt_gamma1[1, 2]))
  lrt_gamma1[, 5] <- c(NA, stats::pchisq(lrt_gamma1[2, 4], df = lrt_gamma1[2, 3], lower.tail=FALSE))
  
  weibull$p_value[1] <- lrt_gamma1[2,5]
  
  # Generate output
  colnames(logistic) <- c("", "coef", "exp(coef)", "P value")
  colnames(weibull) <- c("", "coef", "exp(coef)", "P value")
  rownames(logistic) <- c("")
  rownames(weibull) <- c("")
  ret<-list(par=opt_model_full$par, value=opt_model_full$value, logistic=logistic, weibull=weibull, lrt_beta1=lrt_beta1, lrt_gamma1=lrt_gamma1) 
  ret
}

lklh.logit.piecew.weib.twoCuts <- function(param, group, time, delta, s, cutpoints){ 
  # Get current values of parameters
  beta0<- param[1]
  beta1 <- param[2]
  gamma0  <- param[3]
  gamma1 <- param[4]
  log_shape1 <- param[5]
  shape1 <- exp(log_shape1)
  log_shape2 <- param[6]
  shape2 <- exp(log_shape2)
  log_shape3 <- param[7]
  shape3 <- exp(log_shape3)
  
  # Linear predictors
  lps1 <- beta0 + beta1*group
  lps2 <- gamma0 + gamma1*group
  
  # Calculate pi(x), f(t|z) and S(t|z)
  pi <- exp(lps1)/(exp(lps1) + 1) 
  surv <- ifelse(time < cutpoints[1], exp(-exp(lps2) * time^shape1),
                 ifelse(time < cutpoints[2], exp(-exp(lps2) * (cutpoints[1]^shape1 + time^shape2 - cutpoints[1]^shape2)),
                        exp(-exp(lps2) * (cutpoints[1]^shape1 + cutpoints[2]^shape2 - cutpoints[1]^shape2 + time^shape3 - cutpoints[2]^shape3))))
  dens <- ifelse(time < cutpoints[1], exp(lps2) * shape1 * time^(shape1-1),
                 ifelse(time < cutpoints[2], exp(lps2) * shape2 * time^(shape2-1),
                        exp(lps2) * shape3 * time^(shape3-1))) * surv
  
  #Calculate the log likelihood
  likterms <- ifelse(dens != 0 & (delta*s)  != 0, log((1 - pi) * dens), 0) +
    ifelse(delta*(1-s) != 0, log(pi), 0) +    
    ifelse((1-delta)*s != 0, log((1 - pi) * surv), 0) +
    ifelse((1-delta)*(1 - s) != 0, log(pi + (1 - pi) * surv), 0)
  
  #Output the negative log likelihood
  -sum(likterms)
}

lklh.model2.ratioTest.beta1.piecew.weib.twoCuts <- function(param, group, time, delta, s, cutpoints){
  # Get current values of parameters
  beta0<- param[1]
  beta1 <- 0
  gamma0  <- param[2]
  gamma1 <- param[3]
  log_shape1 <- param[4]
  shape1 <- exp(log_shape1)
  log_shape2 <- param[5]
  shape2 <- exp(log_shape2)
  log_shape3 <- param[6]
  shape3 <- exp(log_shape3)
  
  # Linear predictors
  lps1 <- beta0 + beta1*group
  lps2 <- gamma0 + gamma1*group
  
  # Calculate pi(x), f(t|z) and S(t|z)
  pi <- exp(lps1)/(exp(lps1) + 1) 
  surv <- ifelse(time < cutpoints[1], exp(-exp(lps2) * time^shape1),
                 ifelse(time < cutpoints[2], exp(-exp(lps2) * (cutpoints[1]^shape1 + time^shape2 - cutpoints[1]^shape2)),
                        exp(-exp(lps2) * (cutpoints[1]^shape1 + cutpoints[2]^shape2 - cutpoints[1]^shape2 + time^shape3 - cutpoints[2]^shape3))))
  dens <- ifelse(time < cutpoints[1], exp(lps2) * shape1 * time^(shape1-1),
                 ifelse(time < cutpoints[2], exp(lps2) * shape2 * time^(shape2-1),
                        exp(lps2) * shape3 * time^(shape3-1))) * surv
  
  #Calculate the log likelihood
  likterms <- ifelse(dens != 0 & (delta*s)  != 0, log((1 - pi) * dens), 0) +
    ifelse(delta*(1-s) != 0, log(pi), 0) +    
    ifelse((1-delta)*s != 0, log((1 - pi) * surv), 0) +
    ifelse((1-delta)*(1 - s) != 0, log(pi + (1 - pi) * surv), 0)
  
  #Output the negative log likelihood
  -sum(likterms)
}

lklh.model2.ratioTest.gamma1.piecew.weib.twoCuts <- function(param, group, time, delta, s, cutpoints){ 
  # Get current values of parameters
  beta0<- param[1]
  beta1 <- param[2]
  gamma0  <- param[3]
  gamma1 <- 0
  log_shape1 <- param[4]
  shape1 <- exp(log_shape1)
  log_shape2 <- param[5]
  shape2 <- exp(log_shape2)
  log_shape3 <- param[6]
  shape3 <- exp(log_shape3)
  
  # Linear predictors
  lps1 <- beta0 + beta1*group
  lps2 <- gamma0 + gamma1*group
  
  # Calculate pi(x), f(t|z) and S(t|z)
  pi <- exp(lps1)/(exp(lps1) + 1) 
  surv <- ifelse(time < cutpoints[1], exp(-exp(lps2) * time^shape1),
                 ifelse(time < cutpoints[2], exp(-exp(lps2) * (cutpoints[1]^shape1 + time^shape2 - cutpoints[1]^shape2)),
                        exp(-exp(lps2) * (cutpoints[1]^shape1 + cutpoints[2]^shape2 - cutpoints[1]^shape2 + time^shape3 - cutpoints[2]^shape3))))
  dens <- ifelse(time < cutpoints[1], exp(lps2) * shape1 * time^(shape1-1),
                 ifelse(time < cutpoints[2], exp(lps2) * shape2 * time^(shape2-1),
                        exp(lps2) * shape3 * time^(shape3-1))) * surv
  
  #Calculate the log likelihood
  likterms <- ifelse(dens != 0 & (delta*s)  != 0, log((1 - pi) * dens), 0) +
    ifelse(delta*(1-s) != 0, log(pi), 0) +    
    ifelse((1-delta)*s != 0, log((1 - pi) * surv), 0) +
    ifelse((1-delta)*(1 - s) != 0, log(pi + (1 - pi) * surv), 0)
  
  #Output the negative log likelihood
  -sum(likterms)
}
```
