## Supplementary material for "Estimating the new event-free survival": Code supplement: est_EFS.R

```
est_EFS <- function(data, time, competing_event, landmark, group = NULL, conf.lev=0.95){
  ## -------------------------------------------------------------------------------------------
  ## Author: Judith Vilsmeier
  ## -------------------------------------------------------------------------------------------
  ## Description: Estimates EFS and computes a log-log transformed confidence interval
  ## -------------------------------------------------------------------------------------------
  ## Required Packages: etm
  ## -------------------------------------------------------------------------------------------
  ## Input:            data: dataset with one row per patient
  ##                   time: name of the column in data which contains the event time points or
  ##                         the time points at which the patients were last observed
  ##        competing_event: name of the column in data which contains the event type in the 
  ##                         competing risks model (1 = treatment failure (recommended to be 
  ##                         recorded on day 1), 2 = EFS-event not recommended to be recorded
  ##                         on day 1 (relapse/death after CR), 0=censored)
  ##               landmark: time point of the landmark
  ##                  group: name of the column in data which contains the group, can be 
  ##                         omitted
  ##               conf.lev: level of the confidence limits, can be omitted
  ## -------------------------------------------------------------------------------------------
  ## Output: A list which includes the event times, the estimated EFS, the landmark time point,
  ##         the value of the estimated CIF_1 at the landmark, the estimated CIF_1 for all                                     
  ##         event time points, the estimated CIF_2 for all event time points, the estimated
  ##         variance of estimated EFS, the level of the confidence limits, the log-log  
  ##         transformed confidence interval, the data in long format used for the etm function
  ##
  ##         If the argument 'group' is omitted the output is only one list described as above,  
  ##         otherwise the output contains several lists as described above (one for each group)
  ## -------------------------------------------------------------------------------------------
  
  if(missing(data)) stop("Argument 'data' is missing with no default")
  if(missing(time)) stop("Argument 'time' is missing with no default")
  if(missing(competing_event)) stop("Argument 'competing_event' is missing with no default")
  if(missing(landmark)) stop("Argument 'landmark' is missing with no default")
  if(!is.data.frame(data)) stop("Argument 'data' must be a data.frame")
  
  call <- match.call()
  time <- with(data,get(call$time))
  competing_event <- with(data,get(call$competing_event))
  if(!is.null(call$group)) group <- as.factor(with(data,get(call$group)))
  
  if(!is.numeric(time)) stop("Argument 'time' must be a numerical vector.") 
  if(!is.numeric(competing_event)) stop("Argument 'competing_event' must be a numerical vector.") 
  if(!all(competing_event%in%c(0,1,2))) stop("Argument 'competing_event' must be a vector with entries 0, 1 or 2.")
  if(all(competing_event%in%c(0,1)) | all(competing_event%in%c(0,2))) stop("Only one event type defined in argument 'competing_event'.")
  if(!is.numeric(landmark)) stop("Argument 'landmark' must be numerical.") 
  if(!(length(landmark) == 1)) stop("Argument 'landmark' must be of length 1")
  if(!is.numeric(conf.lev)) stop("Argument 'conf.lev' must be numerical.") 
  if(conf.lev <= 0 | conf.lev >= 1) stop("Argument 'conf.lev' must be greater than 0 and smaller than 1.")
  
  ## Transition matrix and state.names
  tra <- matrix(ncol=3,nrow=3,FALSE)
  tra[1,2:3] <- TRUE
  state.names <- c("0","1","2")
  
  alpha <- 1-conf.lev
  
  ##### Estimate EFS with AJE in competing risks model #####
  if(is.null(group)){
    data.etm <- data.frame(id=1:dim(data)[1], entry=rep(0,dim(data)[1]), exit=time, from=rep(0,dim(data)[1]), to=ifelse(competing_event==0,"cens",competing_event))
    
    ## Aalen-Johansen estimator in competing risks model
    tr.prob <- etm(data.etm, state.names, tra, "cens", 0)
    times <- unique(c(0, tr.prob$time)) 
    
    ## CIF of treatment failure 
    CIF_1 <- trprob(tr.prob, tr.choice = "0 1", times)
    CIF_1_landmark <- trprob(tr.prob, tr.choice = "0 1", timepoints=landmark)
   
    ## Compute EFS as 1-CIF_1(landmark)-CIF_2(t)
    # CIF of not shifted events (relapse, death), EFS estimated with AJE
    CIF_2 <- trprob(tr.prob, tr.choice = "0 2", times)
    efs <- 1 - CIF_1_landmark - CIF_2
    
    ## Log-log transformed confidence intervals for EFS 
    # Covariance of CIF1 and CIF2 for timepoints t <= landmark is equal to the covariance at t
    cov_bef_landmark <- trcov(tr.prob, c("0 1", "0 2"), timepoints=times[times <= landmark])
    # Covariance of CIF1 and CIF2 for timepoints t > landmark is equal to the covariance at landmark
    cov_aft_landmark <- trcov(tr.prob, c("0 1", "0 2"), timepoints=landmark)
    # Combine covariance for timepoints before "landmark" and timepoints after "landmark" to one vector
    cov <- c(cov_bef_landmark, rep(cov_aft_landmark,length(times)-length(cov_bef_landmark)))
    # Estimate variance of EFS estimated with AJE for all event timepoints
    var <- trcov(tr.prob, tr.choice="0 1", timepoints=landmark) + trcov(tr.prob, tr.choice="0 2", timepoints=times) + cov
    # names(var) <- as.character(times)
    
    # Compute CI (log-log transformed)
    suppressWarnings({
    ci_loglog_upper <- ifelse(!is.na(log(efs)), efs^(exp(qnorm(1 - alpha/2)*sqrt(var)/log(efs))), efs)
    ci_loglog_lower <- ifelse(!is.na(log(efs)), efs^(exp(-qnorm(1 - alpha/2)*sqrt(var)/log(efs))), efs)
    })
    ci_loglog <- rbind(ci_loglog_lower, ci_loglog_upper)
    rownames(ci_loglog) <- c("lower", "upper")
    colnames(ci_loglog) <- as.character(times)
    
    res <- list(times=times, efs=efs, landmark=landmark, CIF_1_landmark=CIF_1_landmark, CIF_1=CIF_1, CIF_2=CIF_2, var=var, conf.lev=conf.lev, ci_loglog=ci_loglog, data=data.etm)
  }else{
    res <- list()
    lev <- levels(group)
    for(i in 1:length(lev)){
      gruppe <- lev[i]
      ind <- group==gruppe
      time_ind <- time[ind]
      competing_event_ind <- competing_event[ind]
      data.etm <- data.frame(id=1:length(time_ind), entry=rep(0,length(time_ind)), exit=time_ind, from=rep(0,length(time_ind)), to=ifelse(competing_event_ind==0,"cens",competing_event_ind), group=gruppe)
      
      ## Aalen-Johansen estimator in competing risks model
      tr.prob <- etm(data.etm, state.names, tra, "cens", 0)
      times <- unique(c(0, tr.prob$time)) 
      
      ## CIF of treatment failure 
      CIF_1 <- trprob(tr.prob, tr.choice = "0 1", times)
      CIF_1_landmark <- trprob(tr.prob, tr.choice = "0 1", timepoints=landmark)
      
      ## Compute EFS as 1-CIF_1(landmark)-CIF_2(t)
      # CIF of not shifted events (relapse, death), EFS estimated with AJE
      CIF_2 <- trprob(tr.prob, tr.choice = "0 2", times)
      efs <- 1 - CIF_1_landmark - CIF_2
      
      ## Log-log transformed confidence intervals for EFS 
      # Covariance of CIF1 and CIF2 for timepoints t <= landmark is equal to the covariance at t
      cov_bef_landmark <- trcov(tr.prob, c("0 1", "0 2"), timepoints=times[times <= landmark])
      # Covariance of CIF1 and CIF2 for timepoints t > landmark is equal to the covariance at landmark
      cov_aft_landmark <- trcov(tr.prob, c("0 1", "0 2"), timepoints=landmark)
      # Combine covariance for timepoints before "landmark" and timepoints after "landmark" to one vector
      cov <- c(cov_bef_landmark, rep(cov_aft_landmark,length(times)-length(cov_bef_landmark)))
      # Estimate variance of EFS estimated with AJE for all event timepoints
      var <- trcov(tr.prob, tr.choice="0 1", timepoints=landmark) + trcov(tr.prob, tr.choice="0 2", timepoints=times) + cov
      
      # Compute CI (log-log transformed)
      suppressWarnings({
      ci_loglog_upper <- ifelse(!is.na(log(efs)), efs^(exp(qnorm(1 - alpha/2)*sqrt(var)/log(efs))), efs)
      ci_loglog_lower <- ifelse(!is.na(log(efs)), efs^(exp(-qnorm(1 - alpha/2)*sqrt(var)/log(efs))), efs)
      })
      ci_loglog <- rbind(ci_loglog_lower, ci_loglog_upper)
      rownames(ci_loglog) <- c("lower", "upper")
      colnames(ci_loglog) <- as.character(times)
      
      if(i == 1){
        res <- list(times=times, efs=efs, landmark=landmark, CIF_1_landmark=CIF_1_landmark, CIF_1=CIF_1, CIF_2=CIF_2, var=var, conf.lev=conf.lev, ci_loglog=ci_loglog, data=data.etm)
      }else{
        res <- list(res,list(times=times, efs=efs, landmark=landmark, CIF_1_landmark=CIF_1_landmark, CIF_1=CIF_1, CIF_2=CIF_2, var=var, conf.lev=conf.lev, ci_loglog=ci_loglog, data=data.etm))
      }
    }
    names(res) <- paste("group", lev)
  }
  class(res)<-"est_EFS"
  return(res)
}
```
