## Supplementary material for "Estimating the new event-free survival": Code supplement: simultaneous_testing.R

```
simul_test_efs <- function(data, time, competing_event, group, landmark, t, conf.lev=0.95){
  ## -------------------------------------------------------------------------------------------
  ## Author: Judith Vilsmeier, based on the R-package 'nph'
  ## -------------------------------------------------------------------------------------------
  ## Description: 
  ## -------------------------------------------------------------------------------------------
  ## Required Packages: multcomp
  ## -------------------------------------------------------------------------------------------
  ## Input:            data: dataset with one row per patient
  ##                   time: name of the column in data which contains the event time points or
  ##                         the time points at which the patients were last observed
  ##        competing_event: name of the column in data which contains the event type in the 
  ##                         competing risks model (1 = treatment failure (recommended to be 
  ##                         recorded on day 1), 2 = EFS-event not recommended to be recorded
  ##                         on day 1 (relapse/death after CR), 0=censored)
  ##                  group: name of the column in data which contains the group
  ##               landmark: time point of the landmark
  ##                      t: time point at which EFS difference should be tested (additionally 
  ##                         to drop at day one)
  ##               conf.lev: level of the confidence limits
  ## -------------------------------------------------------------------------------------------
  ## Output: A list which includes the estimated EFS difference and covariance matrix, a table
  ##         containing the results of the simultaneous testing, the estimated EFS and
  ##         covariance matrices for each group and the level of the confidence limits
  ## -------------------------------------------------------------------------------------------
  
  if(missing(data)) stop("Argument 'data' is missing with no default")
  if(missing(time)) stop("Argument 'time' is missing with no default")
  if(missing(competing_event)) stop("Argument 'competing_event' is missing with no default")
  if(missing(group)) stop("Argument 'group' is missing with no default")
  if(missing(landmark)) stop("Argument 'landmark' is missing with no default")
  if(missing(t)) stop("Argument 't' is missing with no default")
  if(!is.data.frame(data)) stop("Argument 'data' must be a data.frame")
  
  call <- match.call()
  time <- with(data,get(call$time))
  group_factor <- as.factor(with(data,get(call$group)))
  # Transform group to a numerical vector containing 0,1,...
  group <- as.numeric(group_factor)-1 
  competing_event <- with(data,get(call$competing_event))
  
  if(!is.numeric(time)) stop("Argument 'time' must be a numerical vector.") 
  if(!all(time>=1)) stop("Argument 'time' must be in days and cannot be smaller than 1.") 
  if(!all(group%in%c(0,1))) stop("Argument 'group' must only contain two groups.")
  if(all(group==0) | all(group==1)) stop("Only one group defined in argument 'group'.")
  if(!is.numeric(competing_event)) stop("Argument 'competing_event' must be a numerical vector.") 
  if(!all(competing_event%in%c(0,1,2))) stop("Argument 'competing_event' must be a vector with entries 0, 1 or 2.")
  if(all(competing_event%in%c(0,1)) | all(competing_event%in%c(0,2))) stop("Only one event type defined in argument 'competing_event'.")
  if(!is.numeric(landmark)) stop("Argument 'landmark' must be numerical.") 
  if(landmark<1) stop("Argument 'landmark' must be in days and cannot be smaller than 1.")
  if(!is.numeric(t)) stop("Argument 't' must be numerical.") 
  if(t<1) stop("Argument 't' must be in days and cannot be smaller than 1.")
  if(!is.numeric(conf.lev)) stop("Argument 'conf.lev' must be numerical.") 
  if(conf.lev <= 0 | conf.lev >= 1) stop("Argument 'conf.lev' must be greater than 0 and smaller than 1.")
  
  rhs <- c(0,0) 
  
  # Estimate EFS and covariance matrix per group
  for(gruppe in c(0,1)) {
    ind <- group==gruppe
    time_ind <- time[ind]
    competing_event_ind <- competing_event[ind]
   
    # Transition matrix and state.names
    tra <- matrix(ncol=3,nrow=3,FALSE)
    tra[1,2:3] <- TRUE
    state.names <- c("0","1","2")
      
    # Bring data in format for etm function (competing_event = 1 event recommended to be recorded on day 1, competing_event = 2 other EFS-event)
    data.etm <- data.frame(id=1:length(time_ind), entry = rep(0,length(time_ind)), exit=time_ind, from=rep(0,length(time_ind)), to=ifelse(competing_event_ind ==0, "cens", competing_event_ind))
    
    # Estimate CIFs
    tr.prob<- etm(data.etm, state.names, tra, "cens", 0, covariance = T)
    times <- tr.prob$time
    est01 <- trprob(tr.prob, tr.choice = "0 1", timepoints=landmark)
    est02 <- trprob(tr.prob, tr.choice = "0 2", timepoints=t)
    # Estimate EFS at day 1 and day t
    efs_1 <- 1-est01
    efs_t <- 1-est01-est02
    var1 <- trcov(tr.prob, tr.choice="0 1", timepoints=landmark) 
    var2 <- var1 + trcov(tr.prob, tr.choice="0 2", timepoints=t) + 2*trcov(tr.prob, c("0 1", "0 2"), timepoints=min(landmark,t))
    # Covariance matrix
    covar <- var1 + trcov(tr.prob, c("0 1", "0 2"), timepoints=min(landmark,t))
    H <- matrix(c(var1, covar, covar, var2), nrow=2)
    if(gruppe==0){
      V0<-H
      val0<-c(efs_1,efs_t)
    }
    if(gruppe==1){
      V1<-H
      val1<-c(efs_1,efs_t)
    }
  }
  # Compute EFS difference and covariance matrix
  V<-V0+V1
  V<-as.matrix(V)
  values<-val1-val0	
  colnames(V)<-rownames(V)<-names(values)<- c("efs_1", "efs_t")
  obj<-list(est=values,V=V) 
  
  # Test EFS difference at day 1 and day t simultaneously
  glht_res<-glht(obj,alternative = "two.sided",rhs=rhs,coef.=function(object,...){object$est}, vcov.=function(object,...){object$V}) 
  CI<-confint(glht_res,level=0.95)$confint 
  p<-summary(glht_res)$test$pvalues
  
  # Generate table with results
  CI<-as.data.frame(CI)
  CI$p_adjusted<-p
  CI$Time_days<-c(1,t)
  CI$SE<-sqrt(diag(obj$V))
  alpha<-1-conf.lev
  # Unadjusted CI and p-value
  CI$lwr_unadj<-CI$Estimate-qnorm(1-alpha/2)*CI$SE 
  CI$upr_unadj<-CI$Estimate+qnorm(1-alpha/2)*CI$SE 
  CI$p_unadj<-2*(1-pnorm(abs((CI$Estimate-rhs)/CI$SE))) 
  # Bonferroni-Holm
  CI$p_Holm<-p.adjust(CI$p_unadj,"holm")
  CI<-CI[,c(5,1,6:9,2:4,10)]
  names(CI)[names(CI)=="Estimate"]<-paste0("Estimate (group ", levels(group_factor)[2]," - group ", levels(group_factor)[1],")")
  names(CI)[names(CI)=="lwr"]<-"lwr_adjusted"
  names(CI)[names(CI)=="upr"]<-"upr_adjusted"
  rownames(CI)<-1:dim(CI)[1]
  
  ret<-list(est=obj$est,V=obj$V,tab=CI,est0=val0,est1=val1,V0=V0,V1=V1,conf.lev=conf.lev)
  ret
}
```
