## Supplementary material for "Estimating the new event-free survival": Code supplement: CB_efs.R

```
CB_efs <- function(est, times, zeta, n, alpha, t.start, t.end) {
  ## ------------------------------------------------------------------------------------------------
  ## Author: Tobias Bluhmki (function_EP.R), small modifications by Judith Vilsmeier
  ## ------------------------------------------------------------------------------------------------
  ## Description: Computes log-log transformed confidence bands (based on wild bootstrap)
  ## -------------------------------------------------------------------------------------------------
  ## Required Packages: 
  ## -------------------------------------------------------------------------------------------------
  ## Input:     est: EFS estimated on the original dataset
  ##          times: time points at which est changes (event time points)
  ##           zeta: bootstrap data generated from function WB_efs
  ##              n: number of patients
  ##          alpha: nominal level
  ##           noit: number of bootstrap iterations
  ##        t.start: lower margin of the time interval over which the confidence bands are computed 
  ##          t.end: upper margin of the time interval over which the confidence bands are computed 
  ## -------------------------------------------------------------------------------------------------
  ## Output: A table containing the lower and upper limit of the confidence band  
  ## -------------------------------------------------------------------------------------------------
  
  # Added by Judith Vilsmeier
  if(missing(est)) stop("Argument 'est' is missing with no default")
  if(missing(times)) stop("Argument 'times' is missing with no default")
  if(missing(zeta)) stop("Argument 'zeta' is missing with no default")
  if(missing(n)) stop("Argument 'n' is missing with no default")
  if(missing(alpha)) stop("Argument 'alpha' is missing with no default")
  if(missing(t.start)) stop("Argument 't.start' is missing with no default")
  if(missing(t.end)) stop("Argument 't.end' is missing with no default")
  
  if(!is.numeric(est)) stop("Argument 'est' must be a numerical vector.") 
  if(!is.numeric(times)) stop("Argument 'times' must be a numerical vector.") 
  if(!is.matrix(zeta)) stop("Argument 'zeta' must be a matrix")
  if(!is.numeric(n)) stop("Argument 'n' must be numerical.") 
  if(n<0) stop("Argument 'n' cannot be smaller than 0.")
  if(alpha <= 0 | alpha >= 1) stop("Argument 'alpha' must be greater than 0 and smaller than 1.")
  if(!is.numeric(t.start)) stop("Argument 't.start' must be numerical.") 
  if(t.start<0) stop("Argument 't.start' cannot be smaller than 0.")
  if(!is.numeric(t.end)) stop("Argument 't.end' must be numerical.") 
  if(t.end<=0) stop("Argument 't.end' must be greater than 0.")
  if(t.start>=t.end) stop("Argument 't.start' must be smaller than argument 't.end'.")
  
  # Extract all event times between t.start and t.end
  eval.times <- times[times <= t.end & times > t.start]
  
  # Est only for event timepoints between t.start and t.end
  est <- est[times <= t.end & times > t.start]
  
  # Extract number of iterations used in wild bootstrap
  noit <- dim(zeta)[1]
  
  # Start R-Code T. Bluhmki
  # Calculate approximate variance required for g(t)
  var.WB <- apply(zeta, 2, var)
  var.P.approx <- 1/n * var.WB
  
  # Derive weight function g(t)
  g <- ifelse((var.P.approx == 0 | est == 1), 0, (est - 1) * log(1 - est)/sqrt(n * var.P.approx))
  
  # Derive dphi
  dphi <- ifelse((est == 0 | est == 1), 0, 1/((est - 1) * log(1 - est)))
  
  # Derive C.hat
  C.hat <- lapply(1:noit, function(j) {
    g * dphi * zeta[j, ]
  })
  
  # Derive event times between t.start and t.end
  times <- eval.times[eval.times >= t.start & eval.times <= t.end]

  # Derive index of event times between t.start and t.end
  index <- findInterval(times, eval.times)
  
  # Derive supremum of each process C.hat 
  sup.C <- sapply(1:noit, function(it, index) {
    max(abs(C.hat[[it]][index]), na.rm = T)
  }, index = index)
  
  # Derive 1-alpha quantile and transform it
  q.alpha <- as.vector(quantile(sup.C, 1 - alpha))
  d <- ifelse(g == 0, 0, q.alpha/(sqrt(n) * g))
  
  # Computation of confidence band margins
  CB.lower.loglog <- ifelse(index > 0, 1 - (1 - est[index])^(exp(-d[index])), 0)
  CB.upper.loglog <- ifelse(index > 0, 1 - (1 - est[index])^(exp(d[index])),  0)
  
  # Added by Judith Vilsmeier
  # Generate output
  output <- matrix(nrow=length(times), ncol=2)
  rownames(output) <- times
  colnames(output) <- c("lower","upper")
  output[,1] <- CB.lower.loglog
  output[,2] <- CB.upper.loglog
  return(output)
}
```
