## Supplementary material for "Estimating the new event-free survival": Code supplement: CB_linear_diff.R

```
CB_linear_diff <- function(data1, data2, zeta1, zeta2, alpha, t.start, t.end){ 
  ## ------------------------------------------------------------------------------------------
  ## Author: Tobias Bluhmki (function_linear_diff.R), modified by Judith Vilsmeier
  ## ------------------------------------------------------------------------------------------
  ## Description: Derives the linear confidence band for the difference in EFS between groups
  ##              (based on wild bootstrap)
  ## ------------------------------------------------------------------------------------------
  ## Required Packages: etm
  ## ------------------------------------------------------------------------------------------
  ## Input:   data1: data of first group in format for etm function
  ##          data2: data of second group in format for etm function
  ##          zeta1: bootstrap data of first group generated from function WB_efs
  ##          zeta2: bootstrap data of second group generated from function WB_efs 
  ##          alpha: nominal level
  ##        t.start: lower margin of the time interval over which the confidence bands are  
  ##                 computed
  ##          t.end: upper margin of the time interval over which the confidence bands are 
  ##                 computed 
  ## ------------------------------------------------------------------------------------------
  ## Output: A list which includes the unique event times in [t.start,t.end], the estimator of the  
  ##         difference in probabilities and the confidence band margins (lower and upper)
  ## ------------------------------------------------------------------------------------------
  
  # Added by Judith Vilsmeier:
  if(missing(data1)) stop("Argument 'data1' is missing with no default")
  if(missing(data2)) stop("Argument 'data2' is missing with no default")
  if(missing(zeta1)) stop("Argument 'zeta1' is missing with no default")
  if(missing(zeta2)) stop("Argument 'zeta2' is missing with no default")
  if(missing(alpha)) stop("Argument 'alpha' is missing with no default")
  if(missing(t.start)) stop("Argument 't.start' is missing with no default")
  if(missing(t.end)) stop("Argument 't.end' is missing with no default")
  
  if(!is.data.frame(data1)) stop("Argument 'data1' must be a data.frame")
  if(!sum(c("id", "from", "to", "time") %in% names(data1)) != 4) stop("'data1' must contain the right variables for etm function ('id', 'from', 'to', 'time')")
  if(!is.data.frame(data2)) stop("Argument 'data2' must be a data.frame")
  if(!sum(c("id", "from", "to", "time") %in% names(data2)) != 4) stop("'data2' must contain the right variables for etm function ('id', 'from', 'to', 'time')")
  if(!is.matrix(zeta1)) stop("Argument 'zeta1' must be a matrix")
  if(!is.matrix(zeta2)) stop("Argument 'zeta2' must be a matrix")
  if(alpha <= 0 | alpha >= 1) stop("Argument 'alpha' must be greater than 0 and smaller than 1.")
  if(!is.numeric(t.start)) stop("Argument 't.start' must be numerical.") 
  if(t.start<0) stop("Argument 't.start' cannot be smaller than 0.")
  if(!is.numeric(t.end)) stop("Argument 't.end' must be numerical.") 
  if(t.end<=0) stop("Argument 't.end' must be greater than 0.")
  if(t.start>=t.end) stop("Argument 't.start' must be smaller than argument 't.end'.")
  
  # Transition matrix and state.names
  tra <- matrix(ncol=3,nrow=3,FALSE)
  tra[1,2:3] <- TRUE
  state.names <- c("0","1","2")
  
  # Extract number of iterations used in wild bootstrap
  noit <- dim(zeta1)[1]
  
  # Determine number of individuals
  n1 <- length(unique(data1$id))
  n2 <- length(unique(data2$id))
  
  # Estimate CIFs with AJE separately for each group
  P.s.all.1 <- etm(data1, state.names, tra, "cens", s = 0)
  P.s.all.2 <- etm(data2, state.names, tra, "cens", s = 0)
  
  # Extract all event times
  eval.times.1 <- P.s.all.1$time
  eval.times.2 <- P.s.all.2$time
  
  # Added by Judith Vilsmeier:
  # Compute P_{treatment failure}(u_0) = max(P_{treatment failure})
  P_treatFail.1 <- trprob(P.s.all.1, tr.choice = "0 1", eval.times.1) 
  P_treatFail_landmark.1 <- max(P_treatFail.1)
  P_treatFail.2 <- trprob(P.s.all.2, tr.choice = "0 1", eval.times.2) 
  P_treatFail_landmark.2 <- max(P_treatFail.2)
  
  # Determine (sorted) unique timepoints of both groups
  combine.times <- sort(unique(c(eval.times.1, eval.times.2)))
  
  # Derive event times between t.start and t.end
  ana.times <- combine.times[combine.times >= t.start & combine.times <= t.end]
  
  # Include t.start and after t.end in event times if not chosen
  if (ana.times[1] > t.start) {
    ana.times <- c(t.start, ana.times)
  }

  if (tail(ana.times, 1) < t.end) {
    ana.times <- c(ana.times, t.end)
  }
  
  # Select bootstrap processes with lm transition in each divided by respective sample size (cf. (11)) !!
  B.1.predict <- lapply(1:noit, function(j) {
    sapply(findInterval(ana.times, eval.times.1), FUN = function(k) {
      if (k == 0) {
        return(0)
      } else (return(zeta1[j, k]))
    })/sqrt(n1)
  })
  B.2.predict <- lapply(1:noit, function(j) {
    sapply(findInterval(ana.times, eval.times.2), FUN = function(k) {
      if (k == 0) {
        return(0)
      } else (return(zeta2[j, k]))
    })/sqrt(n2)
  })
  
  
  # Determine D.hat in (11) with k(t)=1 !!
  D.hat <- lapply(1:noit, FUN = function(i, B1, B2) {
    abs(B1[[i]] - B2[[i]])
  }, B1 = B.1.predict, B2 = B.2.predict)
  
  # Derive supremum of each process !!
  sup.D <- sapply(1:noit, function(it, index) {
    max(D.hat[[it]], na.rm = T)
  })
  
  # Determine (1-\alpha) quantile !!
  q.alpha <- as.vector(quantile(sup.D, 1 - alpha))
  
  # Estimated difference in probabilities (modified by Judith Vilsmeier)
  efs.1 <- 1 - P_treatFail_landmark.1 - trprob(P.s.all.1, tr.choice = "0 2", ana.times)
  efs.2 <- 1 - P_treatFail_landmark.2 - trprob(P.s.all.2, tr.choice = "0 2", ana.times)
  est <- efs.1 - efs.2
    
  # Derive confidence band margins according to (12) !!
  CB.lower <- est - q.alpha
  CB.upper <- est + q.alpha
  
  #Output
  output <- list(times = ana.times, est = est, CB.lower = CB.lower, CB.upper = CB.upper)
  
  return(output)
}
```
