## Supplementary material for "Estimating the new event-free survival": Code supplement: WB_efs.R

```
WB_efs <- function(data, landmark, noit, s=NULL, tau=NULL){
  ## ---------------------------------------------------------------------------------------
  ## Author: Tobias Bluhmki (function_det_zeta.R), modified by Judith Vilsmeier 
  ## ---------------------------------------------------------------------------------------
  ## Description: Performs the wild bootstrap for EFS estimated by the Aalen-Johansen  
  ##              estimator in the competing risks model
  ## ---------------------------------------------------------------------------------------
  ## Required Packages: etm
  ## ---------------------------------------------------------------------------------------
  ## Input:     data: data in format for etm function
  ##            noit: number of bootstrap iterations
  ##         s & tau: margins of time interval [s,tau] over which the wild bootstrap is  
  ##                  applied, if omitted s is chosen as 0 an tau as the last event  
  ##                  timepoint in data
  ##        landmark: time point of the landmark
  ## ---------------------------------------------------------------------------------------
  ## Output: A matrix in which one row represents one bootstrap iteration. The columns  
  ##         correspond to the event times in [s,tau]
  ## ---------------------------------------------------------------------------------------
  
  # Added by Judith Vilsmeier:
  if(missing(data)) stop("Argument 'data' is missing with no default")
  if(missing(landmark)) stop("Argument 'landmark' is missing with no default")
  if(missing(noit)) stop("Argument 'noit' is missing with no default")
  
  if(!is.data.frame(data)) stop("Argument 'data' must be a data.frame")
  if(!sum(c("id", "from", "to", "time") %in% names(data)) != 4) stop("'data' must contain the right variables for etm function ('id', 'from', 'to', 'time')")
  if(!is.numeric(landmark)) stop("Argument 'landmark' must be numerical.") 
  if(!is.numeric(noit)) stop("Argument 'noit' must be numerical.") 
  if(noit<=0) stop("Argument 'noit' must be greater than 0.")
  
  # Set s to 0 and tau to the last event timepoint in data if they are omitted
  if(is.null(s)) s <- 0
  if(is.null(tau)) tau <- max(data$exit[data$to != "cens"])
  
  if(!is.numeric(s)) stop("Argument 's' must be numerical.") 
  if(s<0) stop("Argument 's' cannot be smaller than 0.")
  if(!is.numeric(tau)) stop("Argument 'tau' must be numerical.") 
  if(tau<=0) stop("Argument 'tau' must be greater than 0.")
  if(s>=tau) stop("Argument 's' must be smaller than argument 'tau'.")

  # Transition matrix and state.names
  tra <- matrix(ncol=3,nrow=3,FALSE)
  tra[1,2:3] <- TRUE
  state.names <- c("0","1","2")
  
  # Start R-Code T. Bluhmki
  # Determine number of individuals
  n <- length(unique(data$id))
  
  # Compute P(s,u) for all event times u 
  P.s.all <- etm(data, state.names, tra, "cens", s = s, t = tau, covariance = F, 
                 delta.na = F)
  
  # Extract all event times in [s,tau]
  eval.times <- P.s.all$time[P.s.all$time <= tau & P.s.all$time > s]
  
  # Compute P(u,*) for all u's in [s,tau]
  P.u.v.all <- lapply(eval.times, FUN = function(v)
  {
    # Determine all times u<=v for summation/integration
    int.times <- eval.times[eval.times <= v]
    # Determine for all u: P(u,v)
    P.u.v.it <- lapply(int.times, FUN = function(u)
    {
      # If u==v then identity matrix
      if (u == v | u == max(eval.times))
      {
        P.u.v <- diag(1, length(state.names))
        rownames(P.u.v) <- state.names
        colnames(P.u.v) <- state.names
        # else etm application
      } else
      {
        P.u.v.etm <- etm(data, state.names, tra, "cens", s = u, t = v, 
                         covariance = F)$est
        # Extract last estimated value
        P.u.v <- P.u.v.etm[, , dim(P.u.v.etm)[3]]
      }
      return(P.u.v)
    })
    names(P.u.v.it) <- as.character(int.times)
    return(P.u.v.it)
  })
  names(P.u.v.all) <- as.character(eval.times)
  
  # Compute zeta for each bootstrap iteration
  zeta <- lapply(1:noit, FUN = function(i) 
  {
    # Derive dxi
    dxi.it <- lapply(1:dim(P.s.all$trans)[1], FUN = function(j)
    {
      return((
        # Only for timepoints where individuals at risk
        ifelse(P.s.all$n.risk[, as.character(P.s.all$trans[j, 1])] == 0, 0,
               # Generation of multipliers G as many as event are observed + summation
               sapply(P.s.all$n.event[as.character(P.s.all$trans[j, 1]), 
                                      as.character(P.s.all$trans[j, 2]), ], FUN = function(l)
                                      {
                                        return(sum(rnorm(l, 0, 1)))
                                      })
               # Divided by the number at risk and multiplying with sqrt(n)
               /P.s.all$n.risk[, as.character(P.s.all$trans[j, 1])])) * sqrt(n))
    })
    names(dxi.it) <- paste(P.s.all$trans$from, P.s.all$trans$to, sep = " ")
    
    #Generate matrix dxi
    dxi.u.all <- lapply(eval.times, FUN = function(u)
    {
      dxi.u <- matrix(0, nrow = length(state.names), length(state.names))
      colnames(dxi.u) <- state.names
      rownames(dxi.u) <- state.names
      
      for(row in 1:dim(P.s.all$trans)[1])
      {
        dxi.u[as.character(P.s.all$trans$from[row]), as.character(P.s.all$trans$to[row])]<- dxi.it[[paste(as.character(P.s.all$trans$from[row]), 
                                                                                                          as.character(P.s.all$trans$to[row]), sep = " ")]][[which(P.s.all$time == u)]]
      }
      # Diagonal elements such that sum of each row euqals 0
      diag(dxi.u) <- (-rowSums(dxi.u))
      return(dxi.u)
    })
    names(dxi.u.all) <- as.character(eval.times)
    
    # Added by Judith Vilsmeier:
    # WB relization of CIF_1(landmark)
    int.times <- eval.times[eval.times <= landmark]
    zeta.1.landmark <- sum(sapply(int.times, FUN = function(u)
    {
      # Matrix product
      summand <- P.s.all$est[, , as.character(u)] %*% dxi.u.all[[as.character(u)]] %*% 
        P.u.v.all[[as.character(max(int.times))]][[as.character(u)]]
      return(summand["0","1"])
    }))
    
    # Modified by Judith Vilsmeier:
    # WB relization of CIF_2(v) for each event time v...
    zeta.it <- sapply(eval.times, FUN = function(v)
    {
      int.times <- eval.times[eval.times <= v]
      # Summation over all times <=v
      zeta.2.v <- sum(sapply(int.times, FUN = function(u)
      {
        # Matrix product
        summand <- P.s.all$est[, , as.character(u)] %*% dxi.u.all[[as.character(u)]] %*% 
          P.u.v.all[[as.character(v)]][[as.character(u)]]
        return(summand["0","2"])
      }))
      return(1 - zeta.1.landmark - zeta.2.v)
    })
    return(zeta.it)
  })
  
  # Generate output matrix
  out <- matrix(0, nrow = noit, ncol = length(eval.times))
  rownames(out) <- as.character(1:noit)
  colnames(out) <- as.character(eval.times)
  for (i in 1:noit)
  {
    out[i, ] <- zeta[[i]]
  }
  return(out)
}
```
