## Supplementary material for "Estimating the new event-free survival": Code supplement: readme.pdf

Code supplement for the manuscript "Estimating the new event-free survival" by J. Vilsmeier, M. Saadati, K. Miah, A. Benner, H. Döhner and J. Beyersmann

For questions, comments or remarks about the code please contact J. Vilsmeier.

The code has been written using R version 4.5.2 (platform x86_64-pc-linux-gnu) with packages multcomp_1.4-29, survival_3.8-3 and etm_1.1.2.
A copy of the output of R’s sessionInfo() can be found on page 2.

The dataset “data_example” contains simulated data that mimic the main features of the real data analysed in the article.

The functions used for the estimation of EFS and the computation of confidence intervals, confidence bands, simultaneous testing and cure models are provided in the subfolder “Functions” and subsequent subfolders (see folder structure below).

The function “simultaneous_testing” is based on the R-package ‘nph’.
The functions “WB_efs”, “CB_efs” and “CB_linear_diff” are based on functions provided in the supplementary code of “A Wild Bootstrap Approach for the Aalen-Johansen Estimator” by Bluhmki et al. (2018) (https://academic.oup.com/biometrics/article/74/3/977/7526051#supplementary-data).

Folder structure:

```
readme.pdf
EFS.R
data_example.RData
/Functions
        est_EFS.R
        ./Cure_model
                log_weib_cure.R
                piecew_log_weib_cure_twoCuts.R
        ./Simultaneous_testing
                simultaneous_testing.R
        ./Wild_bootstrap_and_CB
                CB_efs.R
                CB_linear_diff.R
                WB_efs.R
```

```
> sessionInfo()
R version 4.5.2 (2025-10-31)
Platform: x86_64-pc-linux-gnu
Running under: Ubuntu 22.04.5 LTS

Matrix products: default
BLAS:   /usr/lib/x86_64-linux-gnu/blas/libblas.so.3.10.0
LAPACK: /usr/lib/x86_64-linux-gnu/lapack/liblapack.so.3.10.0  LAPACK version 3.10.0

locale:
 [1] LC_CTYPE=en_US.UTF-8       LC_NUMERIC=C               LC_TIME=de_DE.UTF-8
LC_COLLATE=en_US.UTF-8     LC_MONETARY=de_DE.UTF-8    LC_MESSAGES=en_US.UTF-8
LC_PAPER=de_DE.UTF-8
 [8] LC_NAME=C                  LC_ADDRESS=C               LC_TELEPHONE=C
LC_MEASUREMENT=de_DE.UTF-8 LC_IDENTIFICATION=C

time zone: Europe/Berlin
tzcode source: system (glibc)

attached base packages:
[1] stats     graphics  grDevices utils     datasets  methods   base

other attached packages:
[1] multcomp_1.4-29 TH.data_1.1-3   MASS_7.3-65     mvtnorm_1.3-3   etm_1.1.2
survival_3.8-3

loaded via a namespace (and not attached):
 [1] codetools_0.2-19  Matrix_1.7-4      lattice_0.22-5    splines_4.5.2     zoo_1.8-14
parallel_4.5.2    grid_4.5.2        sandwich_3.1-1    data.table_1.17.0 compiler_4.5.2
tools_4.5.2
[12] Rcpp_1.0.14
```
